# Overweight as a Causal Factor Contributing to Better Survival at the Oldest Old Ages: A Mendelian Randomization Study

**DOI:** 10.1101/2024.05.30.24308211

**Authors:** Hongzhe Duan, Konstantin Arbeev, Rachel Holmes, Olivia Bagley, Deqing Wu, Igor Akushevich, Nicole Schupf, Anatoliy Yashin, Svetlana Ukraintseva

**Author notes:** **Correspondence:** Svetlana Ukraintseva,; Hongzhe Duan.

## Abstract

Overweight, defined by a body mass index (BMI) between 25 and 30, has been associated with enhanced survival among older adults in some studies. However, whether being overweight is causally linked to longevity remains unclear. To investigate this, we conducted a Mendelian randomization (MR) study of lifespan 85+ years, using overweight as an exposure variable and data from the Health and Retirement Study and the Long Life Family Study. An essential aspect of MR involves selecting appropriate single-nucleotide polymorphisms (SNPs) as instrumental variables (IVs). This is challenging due to the limited number of SNP candidates within biologically relevant genes that can satisfy all necessary assumptions and criteria. To address this challenge, we employed a novel strategy of creating additional IVs by pairing SNPs between candidate genes. This strategy allowed us to expand the pool of IV candidates with new ‘composite’ SNPs derived from eight candidate obesity genes. Our study found that being overweight between ages 75 and 85, compared to having a normal weight (BMI 18.5-24.9), significantly contributes to improved survival beyond age 85. Results of this MR study thus support a causal relationship between overweight and longevity in older adults.

## 1 Introduction

Overweight, defined by a body mass index (BMI) between 25 and 30, has been associated with certain health risks, as well as with reduced mortality in older adults (Carr et al. 2023; Chapman 2010; Flegal et al. 2013; Hansel et al. 2015; Johnson and Bales 2014; I. M. Lee et al. 2001; Pes et al. 2019; Porter Starr and Bales 2015; Reaven 2011; Zheng et al. 2021). This phenomenon, sometimes referred to as the “overweight/obesity paradox”, was reported mainly by observational studies; however, it remains unconfirmed whether overweight is *causally* linked to longevity. While observational studies are very valuable for revealing associations between various risk factors and health outcomes, they struggle with unmeasured confounding factors. As a result, uncovering causal relationships may prove challenging. An ideal solution would be well-designed randomized clinical trials (RCTs), where all confounders are evenly distributed between treatment and control groups. Such trials, however, are not readily available for longevity outcomes and are also ethically unsuitable. Fortunately, the wealth of information available in large observational studies can be leveraged by modern causal inference approaches to evaluate the underlying causal relationships between health-related risk factors and outcomes.

One such approach is Mendelian randomization (MR) (Plotnikov and Guggenheim 2019; Wehby et al. 2008), a causal inference method, which capitalizes on the random distribution of genetic variants, allowing the separation of the study population into different groups. If specific alleles are significantly associated with a modifiable risk factor of interest and meet all necessary assumptions and criteria, they can serve as instrumental variables (IVs). These IVs create a setting akin to an RCT and enable researchers to explore causality more effectively. In this study, we applied the MR approach to explore causal relationships between overweight and longevity in participants of the Health and Retirement Study (HRS) and the Long Life Family Study (LLFS). The HRS data were used in primary analysis, and the LLFS data were used for replication purposes.

## 2 Materials and Methods

### 2.1 Data

The HRS is a longitudinal panel study conducted by the University of Michigan and supported by the National Institute on Aging (grant number NIA U01AG009740) and the Social Security Administration. The data collection was launched in 1992. A representative sample of about 20,000 Americans aged 50 years and above is surveyed every two years. The original HRS cohort targeted the population of adults in the contiguous United States born during the years 1931-1941 with a 2:1 oversample of African-American and Hispanic populations. New birth cohorts were added every six years. Data collection includes a mixed mode design combining in-person, telephone, mail, and Internet. The RAND Center for the Study of Aging, with funding and support from the NIA and the Social Security Administration, created easy-to-use longitudinal files for researchers. We used version 2018 RAND data, which includes fourteen waves of core interview data across twenty-six survey years (1992-2018). Consent forms were read and signed by each respondent and collected by the interviewer. More details about the study can be found in Sonnega et al. (2014).

The LLFS is a longitudinal study of exceptional survival, longevity, and healthy aging, which is carried out in four field centers (Boston, New York, Pittsburgh, and Denmark). 4,953 individuals from 539 families of exceptional longevity that are determined by the criteria of Family Longevity Selection Score (FLOSS) ≥7 (Sebastiani et al. 2009) were enrolled into study. The first visit was between 2006 and 2009, and the willing participants completed a second in-person visit during 2014-2017 following the same protocols. Between the visits and after the second visit, participants were continuously contacted annually for telephone follow-up to update vital status, medical history, and general health. More details about LLFS can be found in Wojczynski et al. (2022). We used the March 6, 2023 release of LLFS data provided by the LLFS Data Management and Coordinating Center (DMCC).

For both data sets, ages at death were computed using dates of birth and death. For those who did not die within the follow-up period, ages at censoring were determined from dates of birth and the last follow-up: November 2022 in the LLFS and June 2019 in the HRS data. BMI values were determined by the average of BMI measurements between age 75 and 85. The main characteristics (mean, standard deviation, percentage) of variables for samples used in analyses are presented in Table 1.

**Table 1.** Characteristics of the HRS and LLFS samples used in analyses.

|  | HRS |  |  |  |  | LLFS |  |  |
| --- | --- | --- | --- | --- | --- | --- | --- | --- |
|  | White<br>Males | White<br>Females | Black<br>Males | Black<br>Females | Total | White<br>Males | White<br>Females | Total |
| Continuous variables: mean(SD) |  |  |  |  |  |  |  |  |
| <sup>a</sup>Enrollment Age | 66.67(6.93) | 67.62(6.78) | 65.45(7.41) | 65.66(7.27) | 66.94(6.92) | 77.54(4.58) | 78.18(4.39) | 77.89(4.48) |
| <sup>b</sup>Last Follow-Up Age<br>or age of death | 87.40(5.46) | 89.24(5.62) | 85.90(5.75) | 87.45(6.11) | 88.2(5.68) | 87.63(4.55) | 89.28(4.35) | 88.54(4.51) |
| Binary variables: N(percent) |  |  |  |  |  |  |  |  |
| <sup>c</sup>Education | 1271(91.97) | 1652(94.62) | 103(63.19) | 169(80.86) | 3195(91.29) | 172(90.53) | 203(87.5) | 375(88.86) |
| <sup>d</sup>Ever smoked | 975(70.75) | 776(44.44) | 117(71.78) | 96(45.93) | 1964(56.11) | 116(61.05) | 90(38.79) | 206(48.82) |
| <sup>e</sup>Comorbidity (1) | 924(66.86) | 973(55.73) | 115(70.55) | 138(66.03) | 2150(61.43) | 136(71.58) | 143(61.64) | 279(66.11) |
| <sup>f</sup>Overweight (1) | 807(58.39) | 784(44.90) | 97(59.51) | 124(59.33) | 1812(51.77) | 135(71.05) | 119(51.29) | 254(60.19) |
| <sup>g</sup>Survive ≥85 (1) | 980(70.91) | 1421(81.39) | 100(61.35) | 144(68.90) | 2645(75.57) | 146(76.84) | 204(87.93) | 350(82.94) |

Notes: Abbreviations: LLFS – Long Life Family Study, March 6, 2023 release; HRS – Health and Retirement Study, RAND Longitudinal File 2018 (v1); SD – standard deviation. The numbers in table are the numbers of participants used in analysis. Binary variables are coded as: Overweight between age 75-85: 1 – overweight, 0 – normal weight; Survive ≥85: 1 – life span ≥ 85 years, 0 – died before age 85; Comorbidity (presence of cancer, diabetes or cardiovascular diseases): 1 – yes, 0 - no; Education: 1 - High school or above, 0 – below high school; Ever smoked: 1 – yes, 0 - no. Number of people with missing values: LLFS: <sup>a</sup>: 10, <sup>b</sup>: 10, <sup>c</sup>: 26, <sup>d</sup>: 47, <sup>e</sup>: 13, <sup>f</sup>: 199, <sup>g</sup>: 10, HRS: <sup>a</sup>: 1, <sup>b</sup>: 1, <sup>c</sup>: 45, <sup>d</sup>: 98, <sup>e</sup>: 0, <sup>f</sup>: 22, <sup>g</sup>: 1.

### 2.2 Genotyping and candidate genes

Genetic data on 15,620 HRS respondents were provided by the database of Genotypes and Phenotypes (dbGaP), dbGaP Study Accession: phs000428.v2.p2. Genotyping was performed by the National Institute of Health (NIH) Center for Inherited Disease Research (CIDR) (see details in Sonnega et al. (2014)). The HRS used Illumina’s Human Omni2.5-Quad (Omni2.5) BeadChip to genotype 2.4 million single nucleotide polymorphisms (SNPs). The LLFS used similar genotyping platform. 4692 LLFS participants have genotyping information in our data. Blood samples were processed at University of Minnesota and genotyping was performed by the CIDR. Details on genotyping in LLFS are provided in Lee et al. (2013).

Mendelian Randomization can offer robust causal inferences provided that genetic variants used as IVs have plausible biological links with the risk factor (Burgess et al. 2018). We, therefore, selected SNPs in eight obesity/overweight related genes that were reported in the literature (Choquet and Meyre 2011; Walley et al. 2009) as candidates for constructing the IVs in our MR study: ADIPOQ, FTO, LEP, LEPR, INSIG2, MC4R, PCSK1, and PPARG. We aimed to ensure a stronger association between SNPs and the exposure of interest, as SNPs in the obesity related genes tend to be correlated with increased BMI/weight. Table 1s (Supplementary Material) shows the numbers of SNPs available in each gene after Quality Control (QC) in HRS and LLFS data, as well as the locus and functional descriptions, according to National Center for Biotechnology Information Reference Sequence Database. QC procedures were performed based on published protocols (Anderson et al. 2010; Marees et al. 2018).

### 2.3 Instrumental variables

The validity of MR method depends heavily on several key assumptions (Greenland 2018). This may result in a bottle neck scenario where a majority of the IV candidates are discarded due to their failure to satisfy all requirements. Due to limited number of individual genetic variants in candidate obesity genes, we introduced a novel method to create additional ‘composite’ SNPs, and therefore substantially expanding the pool of candidate IVs (see details in section 2.7 “‘Composite’ SNPs creation”). We then selected qualified IVs for further downstream MR analysis.

### 2.4 Assumptions

For a genetic variant to be eligible as an instrumental variable, it is critical that it satisfies three key assumptions (Greenland 2018) (Figure 1).

**Figure 1:**
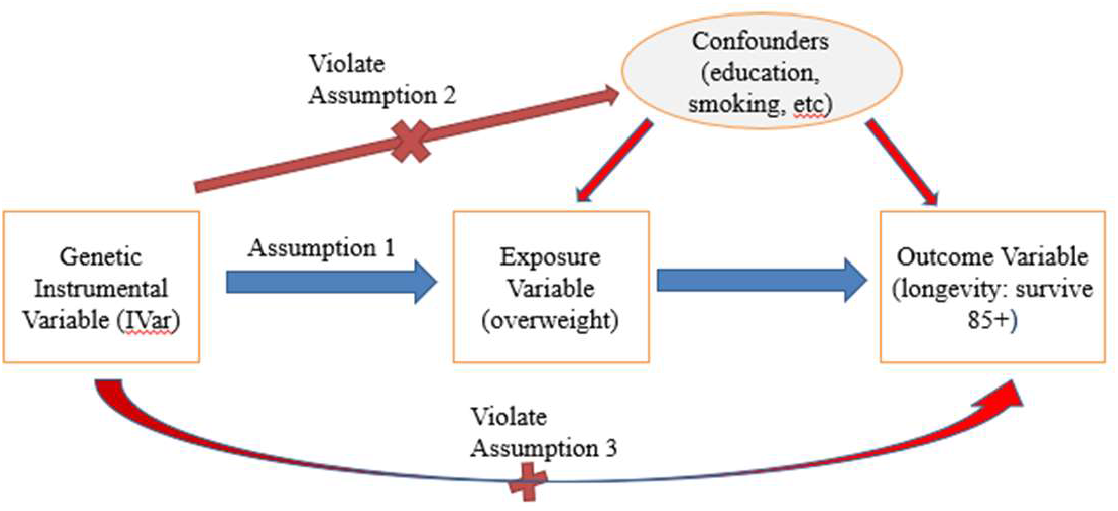
Instrumental variable assumptions in MR. Note: Red lines with crosses denote violations of assumptions if significant associations were identified. Blue line denotes that significant association should exist.

1. Relevance assumption: The SNP must be associated with the exposure of interest.
2. Independence assumption: The SNP is not associated with any confounders.
3. Exclusion assumption: The SNP should be independent of the outcome given the exposure and confounders.

These assumptions can be expressed in the following equations (1) – (3) respectively:

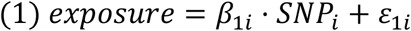

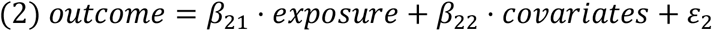

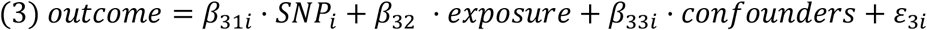

Here coefficient *β*_1*i*_ should be statistically significant (see Strength of instrumental variables section for detail), i.e., SNP has to be strongly associated with the exposure. Covariates with significant *β*_22_ are identified as confounders, and we then test their associations with each SNP. Any SNP associated with any of the identified confounders is removed. Coefficient *β*_31i_ should not be significant, i.e., an IV should not be associated with the outcome with the presence of exposure and confounders because an IV should influence the outcome only through the effect of exposure (Figure 1).

Since our exposure variable is dichotomized from a continuous variable, BMI, we also tested the continuous variable BMI measured during the same age period of interest (see description of variables below in **Analysis**), i.e., between age 75 and 85, for these assumptions (Burgess and Labrecque 2018). All selected genetic variants passed both sets of tests, i.e., for both dichotomized and continuous BMI.

### 2.5 Independence between SNPs

The dependence between SNPs is a potential source of violation for assumption 3, as an IV could influence the outcome through its effect on another IV. To avoid this issue, we calculated the coefficient of determination (*r*^2^) for each SNP pair based on their genotype allele counts to determine their correlations. We used a pre-defined threshold of 0.3 as cutoff point for *r*^2^. If correlation is measured between any pair of SNPs, the SNP least associated with exposure variable is removed. Table 1s shows the starting count of SNPs in each gene after quality control procedures.

### 2.6 Strength of instrumental variables

To ensure the strength of selected IVs, we applied criterion of *F* value > 10 (Burgess et al. 2013) to confirm selected SNPs are associated with exposure with enough significance (assumption 1). The *F* value can be calculated by the following formula (Palmer et al. 2012):

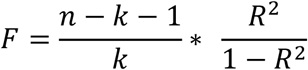

where *n* is sample size, *k* denotes given number of SNPs (here is *k* = 1 since we calculated this value for each SNP), and *R*^2^ denotes the coefficient of determination, which is the proportion of variation explained by the SNP.

In unrelated samples, standard logistic regression is employed. The maximum rescaled R square (Nagelkerke 1991) was calculated by dividing the regular R square by its upper bound to address when the upper bound less than 1. *F* values were calculated using the formula above, with *F* value > 10 considered as fulfillment of this criterion. However, in the case of related samples, such as in LLFS, a mixed model was chosen for analyses, which has an unclear definition of R square. As such, we used a slightly different strategy. First, we calculated residuals as alternative outcomes using SAS GLIMMIX procedure, and then use a general linear model to regress the SNP on the residuals. R squares were taken and *F* values were calculated using the above formula.

### 2.7 ‘Composite’ SNPs creation

When testing SNPs from candidate genes, few, if any of the original SNPs successfully pass all required tests. This issue becomes a significant bottleneck for reliable MR analysis, since the validity of the causal effect identified by MR analysis heavily depends on the qualified IVs that satisfy all assumptions. To address this issue, we tested the effect of SNP-SNP interaction on the exposure variable using SNPs from the same obesity/BMI related genes. We found that numerous SNP pairs are significantly associated with the exposure (Figure 1s), which suggests that we can create a ‘composite’ SNP by pairing the candidate SNPs to expand the candidate pool for IVs.

Our approach involved first selecting all original SNPs that passed quality control from candidate genes and pairing them with each other. Then we summed the dosages of minor alleles (MA) of the two original SNPs and used it to denote the new MA count of the paired SNPs (see Figure 2).

**Figure 2:**
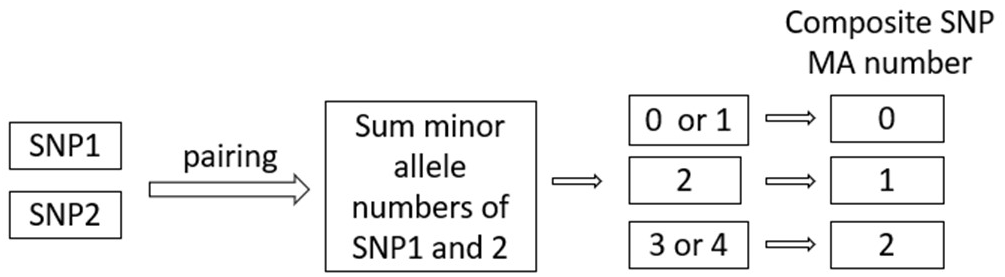
Determination of MA number of ‘composite’ SNP.

For each new pair of SNPs, we denote its MA as follows: 0 if the sum is 0 or 1, 1 if the sum is 2, and 2 if the sum is 3 or 4. In the subsequent analysis, we treated these ‘composite’ SNPs as ‘original’ SNPs with newly denoted minor allele numbers. We then created new binary Plink genetic data incorporating these new ‘SNP’s, and tested all three key assumptions, as well as *F* value and LD criteria. MR analysis was then performed using those ‘composite’ SNPs that survived all of these tests. To avoid confusion, we will use the term ‘composite SNPs’ for the paired SNPs, and ‘single SNPs’ for the original unpaired SNPs in the remainder of this paper.

### 2.8 Analysis

Table 1 describes the key characteristics of study populations of HRS and LLFS data. Our study adhered to the STROBE-MR (Skrivankova et al. 2021) reporting guidelines (checklist is provided in the Supplementary Material Table 2s). For each dataset, we created survival outcome variable (group 1: survived age 85 or above; group 0: died before age 85), and exposure variable (group 1: average BMI ≥ 25 and < 30 (“overweight”) at ages [75,85]; group 0: average BMI ≥ 18.5 and < 25 (“normal weight”) at ages [75,85]). Covariates used include sex (1 – male, 2 – female), race (1 – white, 2 – black, 3 - others), education (0 – below high school, 1 – high school, 2 – above high school), smoking status (1 – ever smoked, 0 – never smoked), first two principal components, and comorbidity (presence of cancer, diabetes or cardiovascular disease (CVD), 1 – yes, 0 - no). For LLFS data, we also included field center (1 – US, 2 – Denmark) as a covariate. Individuals with any missing value(s) were excluded from the study. We did not use covariates in calculating statistics from composite SNP-risk factor and composite SNP-outcome associations for downstream MR analysis (Hartwig et al. 2021).

**Table 2.** MR analysis results in HRS data using composite SNP from obesity/BMI related genes as IV.

| strata | No. of<br>Indiv. | No.<br>of<br>IVs | IVW<br>Estimate<br>(95% CI) | IVW<br>P value | Weighted<br>Median<br>Estimate<br>(95% CI) | Weighted<br>Median<br>P value | MR-Egger<br>Regression<br>Intercept<br>(95%CI) | E-value<br>Average(min,<br>max) |
| --- | --- | --- | --- | --- | --- | --- | --- | --- |
| White<br>female | 1746 | 52 | 0.189<br>(0.031,0.404) | **5.926E-3** | 0.214<br>(0.026,0.402) | **2.541E-2** | 0.044<br>**(-0.060,0.147)** | 2.14<br>(1.60,3.35) |
| White<br>male | 1382 | 51 | 0.158<br>(0.041,0.275) | **8.226E-3** | 0.188<br>(0.021,0.354) | **2.713E-2** | -0.074<br>**(-0.175,0.028)** | 2.29<br>(1.65,3.67) |
| White<br>F+M | 3128 | 69 | -0.00032<br>(-0.108,0.108) | 9.95E-1 | -0.00052<br>(-0.155,0.154) | 9.95E-1 | -0.025<br>(-0.079,0.029) | 2.03<br>(1.46,3.73) |
| Black<br>female | 209 | 144 | 0.057<br>(-0.119,0.312) | 1.151E-1 | 0.034<br>(-0.117,0.481) | 5.137E-1 | -0.075<br>(-0.231,0.081) | 3.23<br>(2.24,9.19) |
| Black<br>male | 163 | 76 | 0.115<br>(0.022,0.207) | **1.529E-2** | 0.108<br>(-0.025,0.241) | 1.122E-1 | 0.047<br>**(-0.210,0.305)** | 3.15<br>(2.39,7.06) |
| Black<br>F+M | 372 | 175 | 0.144<br>(0.083,0.205) | **3.502E-6** | 0.130<br>(0.044,0.216) | **3.119E-3** | -0.057<br>**(-0.150,0.037)** | 3.02<br>(1.92,8.68) |

Coefficients, *β*_Xj_ and *β*_Yj_, along with standard errors were calculated using logistic regression models (4) and (5) due to the binary nature of both the exposure and outcome variable:

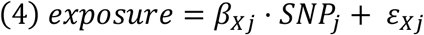

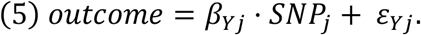

SAS 9.4 was used to calculate these statistics, which were then delivered to R package (‘MendelianRandomization’) for downstream MR analysis adjusted for association between outcome and risk factor to address the fact that we applied one-sample MR analysis in this study.

The Inverse-Variance Weighted (IVW) method, widely used in health studies, was used to evaluate causal estimates (Burgess and Thompson 2017). It benefits from an explicit expression, averaging the ratio estimates from each IV using an inverse-variance weighted formula and provides an overall causal estimate. The causal estimate of IVW is averaged *β*_Yj_/*β*_Xj_ of all IVs. Significant was determined based on having statistically significant IVW, and MR-Egger regression intercept non-distinct from the origin. P value < 0.05 was considered statistically significant.

IVW is a commonly used approach, but suffers from bias if all IVs are not valid. To address this issue, we employed a weighted median approach (Bowden et al. 2016) as a complementary analysis. Ratio estimates of each SNP are ordered and weighted by the inverse of variance. The median MR estimate is considered unbiased if at least 50% of the total weight comes from valid IVs, therefore, it is rather robust. This approach assumes no single IV can contribute more than 50% of the weight.

Additionally, we computed the Kaplan-Meier estimates of survival curves (Figure 2s) to visually illustrate the survival difference between the overweight and normal weight groups. Age at baseline or age 75 years (whichever was the largest) was used as the left truncation variable.

### 2.9 Sensitivity analysis

Sensitivity analyses were performed to assess the sensitivity of specific IVs sets using SNPs obtained from genome-wide association study (GWAS) conducted on LLFS data. We performed a GWAS using the same dichotomized exposure variable (group 1: overweight, at ages [75,85]; group 0: normal weight, at ages [75,85]) as the trait, and the same covariates described in Analysis section. All SNPs with p-value < 0.05 from the GWAS were considered as IV candidates. Then, those that satisfied all key assumptions and passed LD and *F* value criteria were selected as IVs to perform MR analysis. To address relatedness between samples, generalized mixed model (SAS GLIMMIX procedure) was used to calculate statistics from formulae (4) and (5). Results are shown in Table 4s. In this analysis, we used penalized IVW method if standard IVW method did not show significance to address the issue of relatively weaker IV in LLFS data (Xu et al. 2023).

To address the concern that residual unmeasured confounding may compromise causal induction in this observational study, E-values were computed. The *E*-value is defined as the minimum strength of association on the risk ratio (RR) scale that an unmeasured confounder would need to have with both the exposure and the outcome, conditional on the measured covariates, to completely explain away an observed exposure–outcome association. This helps rule out spurious association even with statistically significant results. For each IV, we used the following formula to calculate the E-value (Swanson and VanderWeele 2020).

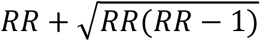

### 2.10 Pleiotropy assessment

In addition to assumptions test described above, we also employed MR-Egger regression (Bowden et al. 2015) to address the issue of potential pleiotropy. SNPs from obesity/BMI related genes often involve different mechanisms, and in turn impact longevity in various ways. These pleiotropy effects are difficult to assess directly. Despite using a predefined small LD to remove SNPs that are correlated with others, residual correlation may still exist to some extent. The intercept of MR-Egger regression offers unbiased evidence for pleiotropy effect. If the regression intercept is observed as non-distinct from the origin, it provides confidence that pleiotropy does not bias the causal effect.

## 3 Results

### 3.1 Descriptive Analyses

Table 1 shows the characteristics of the HRS and LLFS samples at the baseline visit and follow-up. The number of missing values for each variable can be found in the notes under the table. The numbers shown in the table are derived from the participants that were selected in this analysis. Participants need to have a valid value for exposure and outcome variables, as well as all other covariates, to be included in the study. Furthermore, they must have mean BMI measurements ranging from 18.5 to 30 (including normal and over-weight) during the age of between 75 and 85 years old.

Comparatively, the age at enrollment is about 22 years older in LLFS than in HRS, which represents the specific selection of exceptional longevity families in LLFS. HRS participants exhibit a higher proportion of highly educated people and a history of smoking, but a lower incidence of major diseases, including diabetes, cancer, and cardiovascular diseases. Notably, this difference can be explained by different age ranges in the two samples. When we compare the two cohorts at specific age range 75-85, 56.18% of LLFS participants have the above major diseases, compared with 60.16% in HRS data. Moreover, LLFS has a relatively late onset of the above major diseases (67.95 vs 65.33 in HRS) at the same age range. The ages at death or last follow-up are similar in the two data sets. Furthermore, we observed a higher percentage of participants survived beyond 85 years old in LLFS, along with a higher percentage of overweight between ages 75 and 85, as expected.

### 3.2 Mendelian Randomization

Associations between genetic variants and exposure, as well as between genetic variants and outcome are reported (Table 3s 1-6 for HRS, 7-8 for LLFS). Stratified and non-stratified MR results including causal effect estimates, 95% confidence intervals and P-values using HRS and LLFS data, are reported in Table 2 and 3, respectively. In general, a positive IVW effect estimate suggests that an increase in the exposure is associated with an increase in the outcome. In our study, this means overweight at age 75-85 increase the chance of surviving above age 85. The larger estimate implies that the exposure has a more substantial impact on the outcome, assuming all other factors remain constant.

**Table 3.** MR analysis results in LLFS data using composite SNP from obesity/BMI related genes as IV.

| strata | No. of<br>Indiv. | No.<br>of<br>IVs | IVW<br>Estimate<br>(95% CI) | IVW<br>P value | Weighted<br>Median<br>Estimate<br>(95% CI) | Weighted<br>Median<br>P value | MR-Egger<br>Regression<br>Intercept<br>(95%CI) | E-value<br>Average(min,<br>max) |
| --- | --- | --- | --- | --- | --- | --- | --- | --- |
| White<br>female | 232 | - | - | - | - | - | - | - |
| White<br>male | 190 | 135 | 0.192<br>(0.145,0.239) | **1.016E-15** | 0.243<br>(0.180,0.307) | **6.407E-14** | **0.442**<br>**(-0.088,0.795)** | **5.26**<br>**(3.78,6.61)** |
| White<br>F+M | 422 | 47 | 0.307<br>(0.206,0.408) | **2.517E-9** | 0.296<br>(0.164,0.428) | **1.069E-5** | -2.632<br>(-5.115,-0.149) | 4.37<br>(3.67,4.68) |

In HRS data, MR analysis revealed that being overweight at ages 75-85 had a significant causal effect on improved survival above age 85, compared with normal weight at this age range, in white female (IVW p-value=5.926E-3), white male (IVW effect estimate=0.189, p-value=8.226E-3), black male (IVW effect estimate=0.115, p-value=1.529E-2), and non-stratified black subsamples (IVW effect estimate=0.144, p-value=3.502E-6). The most significant results were observed in non-stratified black samples (Table 2). However, the same significant level of causal effect was not seen in black females, warranting further evaluation. All significant results are supported by a significant weighted median test (except for black males) and have survived the MR-Egger regression intercept test.

In LLFS data (Table 3), being overweight at ages 75-85 showed protective effect on living beyond age 85 in white males (IVW effect estimate=0.192, p-value=1.016E-15), but the same effect was not seen in white females as none of the composite SNPs met all criteria. However, a similar effect was observed in white non-stratified samples (IVW effect estimate=0.307, p-value=2.517E-9); weighted median tests are also significant for these two tests. Non-stratified samples did not pass MR-Egger regression intercept test though.

In a sensitivity analysis using LLFS data, we selected the single SNPs significantly associated with exposure variable (p<0.05) from GWAS results. All SNPs satisfied each assumption and criteria as IVs. We observed significant causal effect on life span over 85 years old in penalized IVW method and weighted median test in male (penalized IVW effect estimate=0.156, p-value=7.905E-3, weighted median p-value=6.203E-3) and non-stratified participants (penalized IVW effect estimate=-0.2, p-value=2.142E-3, weighted median p-value=3.401E-3). The standard IVW did not show significance. Both results (Table 4s) and statistics (Table 3s 9-11) are reported.

In another sensitivity analysis, we calculated E-values to test the robustness of our causal effect estimation to residual confounding (Tables 2, 3). Although there were no universally accepted or standardized ad-hoc thresholds for the E-value in the context of Mendelian randomization, an *E*-value of 2 or 3 is considered large enough to reasonably conclude that any residual confounding is unlikely to explain away the causal effect.

## 4 Discussion

Our MR study found that being overweight between the ages 75-85 years significantly contributes to a better survival at ages 85 and beyond in both HRS and LLFS participants. An earlier MR analysis utilizing the UK Biobank data found that the significance of a high BMI as a risk factor in coronary artery disease (CAD) declines in older age (Jansen et al. 2022). Other studies suggested that being moderately overweight could be a marker of a healthy aging that may also protect, at least in part, against comorbidities (Chapman 2010; I. M. Lee et al. 2001; Pes et al. 2019; Porter Starr and Bales 2015; Zheng et al. 2021). Our MR findings are broadly in line with these earlier studies and strongly support the idea that being overweight is a causal factor for longevity. This can have an important implication for clinical practice.

Physicians often consider overweight, defined by BMI between 25 and 30, as detrimental to health, and commonly recommend lifestyle changes to decrease BMI. While some major conditions, such as cardiovascular diseases and type II diabetes, have been associated with high BMI, our study found that BMI that is moderately higher than ‘normal’ (18.5-24.9) could be beneficial for lowering all-cause mortality risk in the very old. Potential mechanisms may involve improved resilience of overweight individuals to late-life stressors (Cho et al. 2018; de Miguel-Diez et al. 2022; Nie et al. 2014; Prescott and Chang 2018). Indeed, larger energy reserves in overweight individuals may be essential for recovery after adverse health events (e.g., pneumonia, fractures), treatments, or surgeries, commonly experienced at the oldest old ages. Conversely, lower energy reserves may adversely affect the capacity of the immune system to fight infections and address harmful exposures. Additional amounts of fat may also better protect older individuals from fall-related fractures, which is a leading factor contributing to mortality in advanced years of life. Altogether, this suggests that attempts to lose weight (e.g., via diet or meds) in excess to its natural aging-related decline may reduce resilience and increase vulnerability to death in the very old. Additional MR studies conducted in different datasets may help further clarify these mechanisms.

Our results also imply that overweight may differently influence longevity in males and females, indicating that they should be analyzed separately. Analyses of combined and unstratified samples yielded less significant and consistent results. For instance, in LLFS, sensitivity analysis found an adverse effect of overweight on survival, while in HRS it showed protective effect, without statistical significance. This discrepancy between the data might result from different study designs. E.g., LLFS cohort has a shorter follow-up period with only two visits, while HRS has fourteen waves of core interviews. This fact could potentially affect calculated mean weight.

Several other factors may also contribute to the discrepant results between the HRS and LLFS data. For example, although BMI is widely used as a criterion of obesity, it is difficult to differentiate between the amount of fat and the amount of muscle. Additionally, studies have shown a significant pattern of assortative mating for BMI (Conley 2016), resulting in the clustering of body weight. These factors are likely more significant in the LLFS data and could act as effective confounders. It is worth noting that SNPs from the eight candidate genes may have different biological effects on obesity and overweight, despite the fact that these genes are all significantly correlated with these traits.

In this paper, we introduced a novel method for constructing and selecting instrumental variables for MR studies. According to our search of up-to-date literature, this is the first time this method has been applied for such purpose. Our approach offers several advantages. Firstly, it uses a straightforward computation and can be easily performed by other researchers. Secondly, it greatly increases the number of the IV candidates and is more likely to lead to a successful selection of qualified IVs. We used all the qualified ‘composite’ SNPs as multiple IVs for the downstream analysis. Alternatively, these IVs can be combined into a ‘polygenic risk score’ by summing their weighted effect alleles, and using this score as a single IV in downstream MR analysis (Dudbridge 2021). However, careful consideration of assumptions and potential biases is crucial when making the decision to apply such score as single IV. Our strategy of leveraging the SNPxSNP interactions to construct IVs from the single SNPs selected from biologically relevant genes was successful and we plan to expand the list of the candidate genes in future analyses.

We acknowledge several study limitations. We did not take advantage of HRS sample weights, limiting us from making inferences at the population-level. This is due to the use of a subsample of HRS data, and binary variables based on the continuous measurement of BMI. Additionally, we analyzed a unique sample (LLFS) selected for exceptional longevity (which was the goal of LLFS); the LLFS participants also have better health and function in several domains compared to other cohorts. Therefore, the results are not generalizable to the general population. Furthermore, limited number of participants in LLFS data decreased the power of this analysis to detect the underlying causal relationship. The relatively small number of LLFS participants increased the difficulty of identifying appropriate SNPs as IVs as the *F* value changes almost linearly with sample size. We also assumed that the genetic variants exhibit consistent associations with the outcome across various strata, such as different age groups and ethnicities, which could affect result interpretation, especially in unstratified analysis. We didn’t investigate other potential confounders (such as gait speed, or other physical activity measures) beyond the ones listed as covariate. The potential issues of pleiotropy were addressed by intercept testing of MR-Egger regression.

## Author Contributions

SU conceived and designed the study, supervised statistical analyses and data preparations, provided interpretation of the findings, and wrote the manuscript. HD prepared data, designed the study, performed statistical analysis, contributed to interpretation of the findings, and wrote the manuscript. RH contributed to programming, proofreading, and writing the manuscript. OB contributed to data preparation. KA, DW, AS, IA, NS, and AY contributed to writing and discussing the manuscript. All authors read and approved the final manuscript.

## Acknowledgments

This study was conducted using data provided by the LLFS, HRS, and dbGaP. We acknowledge researchers and staff who collected these data and made them available for the secondary analyses. The LLFS is sponsored by the National Institute on Aging and carried out in four field centers. The HRS is sponsored by the National Institute on Aging and conducted by the University of Michigan. Access to the HRS genetic data is provided by the database of Genotypes and Phenotypes, dbGaP.

## Ethics Statement

The studies were conducted in accordance with the local legislation and institutional requirements. The studies involving humans were approved by the Duke University Health System Institutional Review Board (IRB). Written informed consents were obtained from the participants in accordance with the LLFS and HRS protocols.

## Data Availability Statement

The LLFS data used in this study were provided by the LLFS Data Management and Coordinating Center (DMCC), Washington University, St. Louis (https://wustl.edu/). The dbGaP also provides access to phenotypic and genetic LLFS data (dbGaP Study Accession: phs000397.v3.p3). The HRS data were provided by dbGaP (Study Accession: phs000428.v2.p2) and the University of Michigan. The authors cannot make data and study materials freely available to other investigators due to dbGaP Data Use Certification Agreement restrictions; however, interested parties can contact NIH dbGaP (https://www.ncbi.nlm.nih.gov/gap/) to request access to dbGaP data through the applicable data access request process. Statistical code is available upon request from the first author.

