## Supplementary Material for "Overweight as a Causal Factor Contributing to Better Survival at the Oldest Old Ages: A Mendelian Randomization Study"

### 1 Supplementary Figures and Tables

**Table 1s**. Description of obesity/overweight related candidate genes selected for this study (the numbers in table show the number of SNPs in each gene after quality control).

| Gene | LLFS | HRS | location | Biological Function and/or relevant process |
|---|---|---|---|---|
| ADIPOQ | 20 | 19 | Chr 3 (186842710..186858463) | Involved with metabolic and hormonal processes. Mutations are associated with adiponectin deficiency. |
| FTO | 345 | 280 | Chr 16 (53703963..54121941) | Studies in mice and humans indicate a role in nervous and cardiovascular systems and a strong association with body mass index, obesity risk, and type 2 diabetes. |
| LEP | 14 | 13 | Chr 7 (128241278..128257629) | Mutations in this gene and its regulatory regions cause severe obesity and morbid obesity with hypogonadism in human patients. |
| LEPR | 113 | 93 | Chr 1 (65420652..65641559) | Mutations have been associated with obesity and pituitary dysfunction. |
| INSIG2 | 15 | 15 | Chr 2 (118088471..118110997) | Block the processing of sterol regulatory element binding proteins. |
| MC4R | 2 | 3 | Chr 18 (60371062..60372775) | Defects in this gene are a cause of autosomal dominant obesity. |
| PCSK1 | 20 | 27 | Chr 5 (96390333..96433248) | Mutations have been associated with susceptibility to obesity and proprotein convertase 1/3 deficiency. |
| PPARG | 122 | 118 | Chr 3 (12287368..12434344) | The protein encoded by this gene is PPAR-gamma, which is associated with obesity, diabetes, atherosclerosis and cancer. |

**Table 2s.** STROBE-MR checklist of recommended items to address in reports of Mendelian randomization studies

| Item No | Section | | Recommendation | Page No |
|---|---|---|---|---|
| 1 | Title and abstract | | Indicate Mendelian randomization (MR) as the study's design in the title and/or the abstract if that is a main purpose of the study | 1 |
| | **INTRODUCTION** | | | |
| 2 | Background | | Explain the scientific background and rationale for the reported study. What is the exposure? Is a potential causal relationship between exposure and outcome plausible? Justify why MR is a helpful method to address the study question | 1 |
| 3 | Objectives | | State specific objectives clearly, including pre-specified causal hypotheses (if any). State that MR is a method that, under specific assumptions, intends to estimate causal effects | 1 |
| | **METHODS** | | | |
| 4 | Study design and data sources | | Present key elements of the study design early in the article. Consider including a table listing sources of data for all phases of the study. For each data source contributing to the analysis, describe the following: | |
| | | a) | Setting: Describe the study design and the underlying population, if possible. Describe the setting, locations, and relevant dates, including periods of recruitment, exposure, follow-up, and data collection, when available. | 2 |
| | | b) | Participants: Give the eligibility criteria, and the sources and methods of selection of participants. Report the sample size, and whether any power or sample size calculations were carried out prior to the main analysis | 2. No power or sample size calculated |
| | | c) | Describe measurement, quality control and selection of genetic variants | 3 |
| | | d) | For each exposure, outcome, and other relevant variables, describe methods of assessment and diagnostic criteria for diseases | 5 |
| | | e) | Provide details of ethics committee approval and participant informed consent, if relevant | 2 |
| 5 | Assumptions | | Explicitly state the three core IV assumptions for the main analysis (relevance, independence and exclusion restriction) as well assumptions for any additional or sensitivity analysis | 3 |
| 6 | Statistical methods: main analysis | | Describe statistical methods and statistics used | |
| | | a) | Describe how quantitative variables were handled in the analyses (i.e., scale, units, model) | 5 |
| | | b) | Describe how genetic variants were handled in the analyses and, if applicable, how their weights were selected | 4-5 |
| | | c) | Describe the MR estimator (e.g. two-stage least squares, Wald ratio) and related statistics. Detail the included covariates and, in case of two-sample MR, whether the same covariate set was used for adjustment in the two samples | 5-6 |
| | | d) | Explain how missing data were addressed | 5 |
| | | e) | If applicable, indicate how multiple testing was addressed | n/a |
| 7 | Assessment of assumptions | | Describe any methods or prior knowledge used to assess the assumptions or justify their validity | 3 |
| 8 | Sensitivity analyses and additional analyses | | Describe any sensitivity analyses or additional analyses performed (e.g. comparison of effect estimates from different approaches, independent replication, bias analytic techniques, validation of instruments, simulations) | 6 |
| 9 | Software and pre-registration | | | |
| | | a) | Name statistical software and package(s), including version and settings used | 5 |

| | | | | |
|---|---|---|---|---|
| | | b) | State whether the study protocol and details were pre-registered (as well as when and where) | n/a |
| | **RESULTS** | | | |
| 10 | Descriptive data | | | |
| | | a) | Report the numbers of individuals at each stage of included studies and reasons for exclusion. Consider use of a flow diagram | Table 1 |
| | | b) | Report summary statistics for phenotypic exposure(s), outcome(s), and other relevant variables (e.g. means, SDs, proportions) | Table 1 |
| | | c) | If the data sources include meta-analyses of previous studies, provide the assessments of heterogeneity across these studies | n/a |
| | | d) | For two-sample MR:<br>i. Provide justification of the similarity of the genetic variant-exposure associations between the exposure and outcome samples<br>ii. Provide information on the number of individuals who overlap between the exposure and outcome studies | n/a |
| 11 | Main results | | | |
| | | a) | Report the associations between genetic variant and exposure, and between genetic variant and outcome, preferably on an interpretable scale | Table 3s |
| | | b) | Report MR estimates of the relationship between exposure and outcome, and the measures of uncertainty from the MR analysis, on an interpretable scale, such as odds ratio or relative risk per SD difference | Table 2, 3 |
| | | c) | If relevant, consider translating estimates of relative risk into absolute risk for a meaningful time period | n/a |
| | | d) | Consider plots to visualize results (e.g. forest plot, scatterplot of associations between genetic variants and outcome versus between genetic variants and exposure) | n/a |
| 12 | Assessment of assumptions | | | |
| | | a) | Report the assessment of the validity of the assumptions | 3 |
| | | b) | Report any additional statistics (e.g., assessments of heterogeneity across genetic variants, such as $I_2$, Q statistic or E-value) | 6 |
| 13 | Sensitivity analyses and additional analyses | | | |
| | | a) | Report any sensitivity analyses to assess the robustness of the main results to violations of the assumptions | 6 |
| | | b) | Report results from other sensitivity analyses or additional analyses | 6 |
| | | c) | Report any assessment of direction of causal relationship (e.g., bidirectional MR) | n/a |
| | | d) | When relevant, report and compare with estimates from non-MR analyses | n/a |
| | | e) | Consider additional plots to visualize results (e.g., leave-one-out analyses) | Figure 1s,2s |
| | **DISCUSSION** | | | |
| 14 | Key results | | Summarize key results with reference to study objectives | 8-9 |
| 15 | Limitations | | Discuss limitations of the study, taking into account the validity of the IV assumptions, other sources of potential bias, and imprecision. Discuss both direction and magnitude of any potential bias and any efforts to address them | 9 |
| 16 | Interpretation | | | |
| | | a) | Meaning: Give a cautious overall interpretation of results in the context of their limitations and in comparison with other studies | 8 |
| | | b) | Mechanism: Discuss underlying biological mechanisms that could drive a potential causal relationship between the investigated exposure and the outcome, and whether the gene-environment equivalence assumption is reasonable. Use causal language carefully, clarifying that IV estimates may provide causal effects only under certain assumptions | 8-9 |

| | | | |
|---|---|---|---|
| | c) | Clinical relevance: Discuss whether the results have clinical or public policy relevance, and to what extent they inform effect sizes of possible interventions | 9 |
| 17 | Generalizability | Discuss the generalizability of the study results (a) to other populations, (b) across other exposure periods/timings, and (c) across other levels of exposure | 9 |
| | **OTHER INFORMATION** | | |
| 18 | Funding | Describe sources of funding and the role of funders in the present study and, if applicable, sources of funding for the databases and original study or studies on which the present study is based | 9 |
| 19 | Data and data sharing | Provide the data used to perform all analyses or report where and how the data can be accessed, and reference these sources in the article. Provide the statistical code needed to reproduce the results in the article, or report whether the code is publicly accessible and if so, where | 9 |
| 20 | Conflicts of Interest | All authors should declare all potential conflicts of interest | 9 |

**Table 3s**. Associations between genetic variants and overweight, and between genetic variants and survive 85+ (pPhenoSNP denotes the p value of association between genetic variants and outcome with the presence of exposure and confounders; pExpSNP denotes the p value of model (5); Beta_1, SE_1 and beta_2 and SE_2 are the coefficients of genetic variants and standard errors for models (4) and (5) respectively; F denotes $F$ value; N denotes number of participants)

Table 3s.1. HRS white females

| SNP | pPhenoSNP | beta_1 | SE_1 | pExpSNP | beta_2 | SE_2 | F |
|---|---|---|---|---|---|---|---|
| rs10490624_rs9922370 | 5.72E-01 | 0.44 | 0.19 | 1.97E-02 | 0.14 | 0.22 | 11.54 |
| rs10493380_rs7194243 | 9.41E-01 | -0.23 | 0.10 | 2.41E-02 | -0.01 | 0.13 | 10.27 |
| rs12629751_rs4135284 | 7.93E-01 | 0.33 | 0.13 | 1.09E-02 | -0.02 | 0.16 | 13.56 |
| rs13428113_rs2665275 | 5.04E-01 | -0.24 | 0.09 | 7.17E-03 | 0.04 | 0.11 | 14.60 |
| rs1559509_rs9302654 | 5.35E-02 | -0.24 | 0.09 | 1.07E-02 | 0.20 | 0.11 | 13.15 |
| rs17036281_rs7206012 | 3.97E-01 | 0.47 | 0.20 | 1.99E-02 | -0.17 | 0.26 | 11.54 |
| rs1801282_rs2042032 | 4.88E-01 | -0.30 | 0.12 | 9.35E-03 | -0.17 | 0.16 | 13.71 |
| rs1805096_rs79497105 | 9.29E-01 | -0.25 | 0.11 | 2.50E-02 | -0.06 | 0.15 | 10.13 |
| rs1887285_rs9922370 | 2.88E-01 | 0.52 | 0.21 | 1.22E-02 | 0.25 | 0.23 | 13.41 |
| rs2028759_rs796313 | 7.51E-01 | -0.68 | 0.27 | 1.12E-02 | -0.14 | 0.32 | 13.76 |
| rs2036373_rs11644943 | 5.97E-01 | 0.36 | 0.14 | 1.20E-02 | -0.02 | 0.18 | 13.14 |
| rs2120825_rs13099634 | 9.19E-01 | -0.42 | 0.17 | 1.48E-02 | -0.10 | 0.23 | 12.01 |
| rs2120825_rs17218700 | 9.26E-01 | -0.41 | 0.17 | 1.88E-02 | -0.04 | 0.22 | 11.32 |
| rs2120825_rs17819063 | 4.60E-01 | -0.42 | 0.17 | 1.51E-02 | -0.27 | 0.24 | 12.09 |
| rs2767485_rs75222427 | 1.43E-01 | -0.58 | 0.20 | 3.26E-03 | -0.58 | 0.31 | 18.20 |
| rs2972165_rs2665275 | 2.91E-01 | -0.37 | 0.15 | 1.30E-02 | -0.17 | 0.20 | 12.55 |
| rs35510800_rs9931580 | 6.80E-01 | 0.22 | 0.08 | 5.01E-03 | 0.06 | 0.10 | 16.11 |

| | | | | | | | |
|---|---|---|---|---|---|---|---|
| rs3792747_rs7192835 | 6.62E-01 | 0.32 | 0.13 | 1.18E-02 | -0.01 | 0.16 | 13.17 |
| rs3792747_rs72807785 | 3.39E-01 | 0.38 | 0.13 | 2.76E-03 | -0.11 | 0.16 | 18.85 |
| rs3792747_rs967515 | 5.81E-01 | 0.32 | 0.13 | 1.69E-02 | 0.13 | 0.16 | 11.84 |
| rs3792747_rs9931580 | 2.16E-01 | 0.24 | 0.09 | 5.86E-03 | -0.09 | 0.11 | 15.56 |
| rs3811942_rs72807785 | 1.61E-01 | 0.30 | 0.12 | 1.02E-02 | -0.17 | 0.15 | 13.66 |
| rs4135263_rs6232 | 3.86E-01 | 0.49 | 0.22 | 2.69E-02 | 0.25 | 0.25 | 10.34 |
| rs4135275_rs11075994 | 5.37E-01 | 0.22 | 0.09 | 1.16E-02 | 0.09 | 0.11 | 13.04 |
| rs4135275_rs16953089 | 2.11E-01 | 0.24 | 0.09 | 5.69E-03 | 0.13 | 0.10 | 15.68 |
| rs4135275_rs17819063 | 4.14E-01 | 0.41 | 0.14 | 3.87E-03 | 0.17 | 0.16 | 17.60 |
| rs4135275_rs35510800 | 6.55E-01 | 0.25 | 0.10 | 9.12E-03 | 0.05 | 0.12 | 13.98 |
| rs4135275_rs3792747 | 1.47E-01 | 0.25 | 0.11 | 2.21E-02 | -0.15 | 0.14 | 10.74 |
| rs4135275_rs4135294 | 4.78E-01 | 0.38 | 0.16 | 1.43E-02 | -0.10 | 0.20 | 12.39 |
| rs4135275_rs708278 | 8.67E-01 | 0.23 | 0.09 | 1.36E-02 | 0.03 | 0.12 | 12.49 |
| rs4135275_rs9922619 | 8.27E-01 | 0.19 | 0.08 | 1.60E-02 | 0.04 | 0.10 | 11.81 |
| rs4135275_rs9931580 | 9.88E-01 | 0.30 | 0.09 | 1.34E-03 | 0.03 | 0.11 | 21.26 |
| rs4135284_rs9922370 | 5.61E-01 | 0.35 | 0.15 | 1.98E-02 | 0.17 | 0.17 | 11.31 |
| rs4783826_rs9924877 | 8.06E-01 | 0.16 | 0.07 | 1.50E-02 | 0.04 | 0.08 | 11.98 |
| rs62034079_rs7206012 | 7.83E-01 | 0.60 | 0.23 | 8.55E-03 | 0.07 | 0.26 | 14.96 |
| rs62034079_rs741300 | 7.38E-01 | 0.22 | 0.09 | 1.88E-02 | -0.01 | 0.12 | 11.29 |
| rs6230_rs77289909 | 9.08E-02 | 0.58 | 0.26 | 2.56E-02 | -0.58 | 0.38 | 10.66 |
| rs6232_rs9925908 | 1.88E-01 | 0.53 | 0.20 | 9.22E-03 | 0.37 | 0.22 | 14.59 |
| rs7185938_rs12051261 | 5.32E-01 | 0.28 | 0.11 | 7.48E-03 | 0.12 | 0.14 | 14.42 |
| rs7186637_rs7206456 | 9.52E-02 | -0.23 | 0.09 | 9.92E-03 | -0.20 | 0.12 | 13.42 |
| rs7190492_rs35510800 | 5.76E-01 | 0.19 | 0.07 | 1.22E-02 | 0.08 | 0.09 | 12.78 |
| rs7200972_rs7194243 | 8.41E-01 | -0.28 | 0.10 | 3.46E-03 | -0.05 | 0.12 | 17.27 |
| rs7206456_rs7194243 | 8.94E-02 | -0.18 | 0.08 | 2.51E-02 | -0.18 | 0.11 | 10.11 |
| rs7626560_rs9934504 | 9.87E-01 | 0.33 | 0.13 | 1.27E-02 | -0.02 | 0.16 | 12.94 |
| rs78663648_rs2120825 | 5.18E-01 | -0.66 | 0.26 | 1.06E-02 | 0.11 | 0.31 | 13.61 |
| rs78663648_rs7194243 | 8.90E-01 | -0.36 | 0.15 | 1.74E-02 | -0.01 | 0.19 | 11.47 |
| rs889904_rs201163647 | 6.82E-02 | -0.16 | 0.06 | 1.21E-02 | -0.17 | 0.08 | 12.71 |
| rs889904_rs2665275 | 3.68E-01 | -0.21 | 0.08 | 1.09E-02 | -0.12 | 0.11 | 13.07 |
| rs889904_rs6233 | 4.93E-01 | -0.15 | 0.06 | 1.84E-02 | -0.06 | 0.08 | 11.21 |
| rs9931580_rs1588413 | 6.45E-01 | 0.21 | 0.09 | 2.37E-02 | 0.09 | 0.11 | 10.47 |
| rs9934504_rs56379708 | 9.10E-01 | 0.26 | 0.11 | 2.08E-02 | -0.05 | 0.14 | 11.00 |
| rs9934528_rs4783826 | 9.01E-01 | 0.17 | 0.06 | 1.03E-02 | 0.05 | 0.08 | 13.32 |

Table 3s.2. HRS white males

| SNP | pPhenoSNP | beta_1 | SE_1 | pExpSNP | beta_2 | SE_2 | F |
|---|---|---|---|---|---|---|---|
| rs1063538_rs11760956 | 8.37E-01 | -0.21 | 0.08 | 5.85E-03 | -0.01 | 0.08 | 16.19 |
| rs1063538_rs79868385 | 5.78E-01 | -0.37 | 0.12 | 2.70E-03 | -0.08 | 0.13 | 19.73 |
| rs1152001_rs3856806 | 5.04E-01 | -0.30 | 0.11 | 6.49E-03 | 0.08 | 0.11 | 16.11 |
| rs1152001_rs60286074 | 9.67E-01 | -0.46 | 0.21 | 2.62E-02 | 0.00 | 0.21 | 11.02 |
| rs1152001_rs79868385 | 5.27E-01 | -0.40 | 0.18 | 2.91E-02 | -0.10 | 0.19 | 10.52 |
| rs1171269_rs6773957 | 6.96E-01 | -0.20 | 0.08 | 1.87E-02 | 0.04 | 0.09 | 11.80 |
| rs12145690_rs7795794 | 5.40E-01 | -0.27 | 0.11 | 1.15E-02 | 0.07 | 0.11 | 13.78 |
| rs12933928_rs1588413 | 4.67E-01 | 0.37 | 0.13 | 3.11E-03 | 0.10 | 0.14 | 18.51 |
| rs12933928_rs9922370 | 5.61E-01 | 0.38 | 0.14 | 8.57E-03 | 0.09 | 0.15 | 14.64 |
| rs1559509_rs1152001 | 3.58E-01 | -0.28 | 0.11 | 1.09E-02 | -0.12 | 0.12 | 14.07 |
| rs16952730_rs1076471 | 7.53E-01 | 0.23 | 0.08 | 4.79E-03 | 0.03 | 0.09 | 16.76 |
| rs16952730_rs7185938 | 7.31E-01 | 0.24 | 0.08 | 3.31E-03 | 0.04 | 0.09 | 18.23 |
| rs17222465_rs9922370 | 4.65E-01 | -0.25 | 0.10 | 1.43E-02 | 0.09 | 0.11 | 12.95 |
| rs17226942_rs7185938 | 2.43E-01 | 0.31 | 0.11 | 7.13E-03 | 0.14 | 0.12 | 15.23 |
| rs17820328_rs918032 | 1.11E-01 | -0.28 | 0.12 | 1.56E-02 | -0.18 | 0.13 | 12.59 |
| rs17823199_rs708262 | 4.98E-01 | 0.18 | 0.08 | 2.37E-02 | -0.07 | 0.09 | 10.75 |
| rs2036373_rs6230 | 7.01E-01 | -0.68 | 0.26 | 7.79E-03 | -0.08 | 0.25 | 16.63 |
| rs2042032_rs12933928 | 4.62E-01 | 0.25 | 0.11 | 2.15E-02 | 0.10 | 0.12 | 11.09 |
| rs2067819_rs17222465 | 6.85E-02 | -0.22 | 0.09 | 1.13E-02 | 0.17 | 0.09 | 13.76 |
| rs2120825_rs7186637 | 6.55E-01 | -0.37 | 0.16 | 1.73E-02 | 0.07 | 0.16 | 12.50 |
| rs3828942_rs2229616 | 8.73E-01 | 0.31 | 0.13 | 1.46E-02 | -0.02 | 0.14 | 12.53 |
| rs3856806_rs12324955 | 2.50E-01 | -0.33 | 0.13 | 1.37E-02 | -0.16 | 0.14 | 13.29 |
| rs3856806_rs28954118 | 6.54E-01 | -0.63 | 0.28 | 2.73E-02 | 0.15 | 0.27 | 11.21 |
| rs4135247_rs17226942 | 4.91E-01 | 0.24 | 0.11 | 2.38E-02 | 0.08 | 0.11 | 10.72 |
| rs4135275_rs41457646 | 3.31E-01 | 0.30 | 0.13 | 1.97E-02 | 0.13 | 0.14 | 11.43 |
| rs4135284_rs9925311 | 7.86E-01 | 0.75 | 0.28 | 6.31E-03 | -0.07 | 0.29 | 16.84 |
| rs41457646_rs1421090 | 1.82E-01 | 0.25 | 0.11 | 2.67E-02 | 0.16 | 0.12 | 10.30 |
| rs41457646_rs1588413 | 8.51E-02 | 0.32 | 0.13 | 1.76E-02 | 0.22 | 0.14 | 11.88 |
| rs41457646_rs4784330 | 4.94E-01 | 0.41 | 0.18 | 2.29E-02 | -0.17 | 0.21 | 10.96 |
| rs4468199_rs79868385 | 3.86E-01 | -0.70 | 0.33 | 3.30E-02 | -0.28 | 0.34 | 10.70 |
| rs4784353_rs1076471 | 5.08E-01 | 0.26 | 0.11 | 2.32E-02 | 0.08 | 0.12 | 10.82 |
| rs62048379_rs967515 | 7.48E-01 | 0.43 | 0.18 | 2.03E-02 | -0.09 | 0.21 | 11.47 |
| rs6230_rs113014601 | 4.99E-01 | -0.57 | 0.26 | 2.81E-02 | -0.20 | 0.27 | 10.80 |
| rs6230_rs12445162 | 8.55E-01 | -0.50 | 0.23 | 2.83E-02 | -0.05 | 0.23 | 10.97 |
| rs6230_rs17820328 | 7.09E-02 | -0.54 | 0.24 | 2.48E-02 | -0.47 | 0.27 | 11.25 |

| | | | | | | | |
|---|---|---|---|---|---|---|---|
| rs6230_rs4783826 | 1.76E-01 | -0.19 | 0.09 | 2.36E-02 | -0.14 | 0.09 | 10.91 |
| rs6230_rs7795794 | 3.91E-01 | -0.57 | 0.22 | 8.57E-03 | -0.16 | 0.22 | 16.02 |
| rs6230_rs79868385 | 1.41E-01 | -0.56 | 0.25 | 2.68E-02 | -0.38 | 0.28 | 11.21 |
| rs7190053_rs57957459 | 6.63E-01 | 0.26 | 0.12 | 2.88E-02 | 0.05 | 0.13 | 10.04 |
| rs822396_rs1063538 | 4.04E-01 | -0.32 | 0.12 | 5.85E-03 | 0.07 | 0.12 | 16.28 |
| rs9436297_rs10749753 | 6.61E-01 | 0.21 | 0.09 | 2.31E-02 | 0.07 | 0.10 | 10.84 |
| rs9436297_rs17415296 | 6.76E-01 | 0.49 | 0.16 | 1.75E-03 | -0.04 | 0.17 | 21.04 |
| rs9436297_rs2003583 | 2.57E-01 | 0.34 | 0.14 | 1.24E-02 | -0.15 | 0.15 | 13.17 |
| rs9436297_rs41457646 | 1.22E-01 | 0.44 | 0.16 | 5.93E-03 | 0.27 | 0.16 | 16.12 |
| rs9436297_rs71390222 | 9.89E-01 | 0.44 | 0.19 | 1.69E-02 | 0.03 | 0.20 | 12.15 |
| rs9436297_rs73136795 | 7.99E-01 | 0.42 | 0.19 | 2.58E-02 | 0.11 | 0.20 | 10.53 |
| rs9436297_rs73617811 | 1.99E-01 | 0.62 | 0.28 | 2.56E-02 | 0.35 | 0.29 | 10.58 |
| rs9436297_rs76442450 | 2.02E-01 | 0.61 | 0.22 | 6.51E-03 | 0.34 | 0.23 | 16.05 |
| rs970468_rs436321 | 5.15E-01 | -0.17 | 0.08 | 2.88E-02 | 0.04 | 0.08 | 10.15 |
| rs9934528_rs77720715 | 5.33E-02 | -0.46 | 0.15 | 2.78E-03 | -0.32 | 0.17 | 19.72 |
| rs9935366_rs17222465 | 8.73E-01 | -0.21 | 0.08 | 8.27E-03 | 0.02 | 0.08 | 14.88 |

Table 3s.3. HRS white non-stratified

| SNP | pPhenoSNP | beta_1 | SE_1 | pExpSNP | beta_2 | SE_2 | F |
|---|---|---|---|---|---|---|---|
| rs1063537_rs62034079 | 7.75E-01 | 0.30 | 0.13 | 2.24E-02 | -0.01 | 0.15 | 10.77 |
| rs11075994_rs9934504 | 7.51E-01 | 0.17 | 0.07 | 2.52E-02 | 0.03 | 0.09 | 10.28 |
| rs11585329_rs2120825 | 3.03E-01 | -0.24 | 0.11 | 2.58E-02 | 0.12 | 0.12 | 10.27 |
| rs1175544_rs7200972 | 2.50E-01 | -0.13 | 0.05 | 7.83E-03 | 0.07 | 0.06 | 14.47 |
| rs12324955_rs7194243 | 3.97E-01 | -0.14 | 0.06 | 2.23E-02 | 0.04 | 0.07 | 10.69 |
| rs12629751_rs4135284 | 4.53E-01 | 0.22 | 0.10 | 1.98E-02 | -0.08 | 0.12 | 11.18 |
| rs13337696_rs9922370 | 7.33E-01 | 0.77 | 0.24 | 1.48E-03 | 0.13 | 0.26 | 22.31 |
| rs13428113_rs2665275 | 9.66E-01 | -0.16 | 0.06 | 1.36E-02 | -0.01 | 0.08 | 12.48 |
| rs1344503_rs7190483 | 2.03E-01 | -0.32 | 0.14 | 2.64E-02 | 0.12 | 0.16 | 10.24 |
| rs1559509_rs9302654 | 1.84E-01 | -0.20 | 0.07 | 4.07E-03 | 0.09 | 0.08 | 16.97 |
| rs16952730_rs9931580 | 7.13E-01 | 0.15 | 0.06 | 1.92E-02 | -0.05 | 0.08 | 11.23 |
| rs17823199_rs708262 | 6.14E-01 | 0.13 | 0.05 | 1.16E-02 | -0.03 | 0.06 | 13.04 |
| rs1801282_rs76330623 | 1.21E-01 | -0.45 | 0.20 | 2.56E-02 | 0.27 | 0.21 | 10.53 |
| rs1805134_rs16952663 | 9.87E-01 | -0.44 | 0.16 | 5.38E-03 | -0.01 | 0.18 | 16.25 |
| rs2003583_rs77720715 | 4.73E-01 | 0.37 | 0.14 | 1.01E-02 | -0.13 | 0.17 | 13.68 |
| rs2028759_rs796313 | 4.53E-01 | -0.61 | 0.20 | 2.68E-03 | 0.12 | 0.23 | 19.23 |

| | | | | | | | |
|---|---|---|---|---|---|---|---|
| rs2071045_rs62034079 | 8.38E-01 | 0.23 | 0.09 | 1.30E-02 | 0.01 | 0.11 | 12.68 |
| rs2120825_rs17218700 | 8.11E-01 | -0.35 | 0.13 | 5.94E-03 | -0.03 | 0.15 | 15.83 |
| rs2120825_rs17819063 | 5.95E-01 | -0.33 | 0.13 | 1.45E-02 | 0.04 | 0.15 | 12.41 |
| rs2120825_rs28954118 | 4.53E-01 | -0.54 | 0.19 | 3.71E-03 | 0.12 | 0.20 | 18.05 |
| rs2120825_rs7195539 | 2.22E-01 | -0.50 | 0.22 | 2.72E-02 | 0.22 | 0.24 | 10.32 |
| rs2882298_rs17823199 | 4.99E-01 | 0.11 | 0.05 | 2.55E-02 | 0.04 | 0.06 | 10.19 |
| rs35808688_rs3856806 | 4.64E-01 | -0.28 | 0.12 | 1.62E-02 | -0.14 | 0.14 | 11.97 |
| rs36219760_rs7795794 | 1.38E-01 | -0.50 | 0.22 | 2.31E-02 | -0.50 | 0.30 | 10.97 |
| rs3751813_rs35510800 | 8.39E-01 | 0.13 | 0.05 | 1.51E-02 | 0.01 | 0.06 | 12.08 |
| rs3751813_rs72807785 | 7.61E-01 | 0.17 | 0.06 | 7.04E-03 | -0.01 | 0.07 | 14.86 |
| rs3856806_rs1558756 | 1.31E-01 | -0.15 | 0.06 | 1.30E-02 | 0.12 | 0.07 | 12.64 |
| rs3856806_rs3826169 | 7.37E-01 | -0.24 | 0.09 | 6.20E-03 | 0.04 | 0.10 | 15.45 |
| rs4135247_rs7194907 | 4.16E-01 | 0.10 | 0.05 | 2.52E-02 | -0.04 | 0.05 | 10.24 |
| rs4135273_rs62034079 | 8.98E-01 | 0.63 | 0.22 | 3.46E-03 | 0.04 | 0.24 | 18.52 |
| rs4135275_rs1076471 | 6.98E-01 | 0.17 | 0.06 | 3.04E-03 | 0.03 | 0.07 | 17.99 |
| rs4135275_rs17819063 | 5.00E-01 | 0.24 | 0.10 | 1.29E-02 | 0.09 | 0.11 | 12.72 |
| rs4135275_rs4135284 | 2.06E-01 | 0.31 | 0.11 | 5.47E-03 | -0.18 | 0.14 | 15.85 |
| rs4135275_rs41457646 | 1.41E-01 | 0.24 | 0.09 | 9.03E-03 | 0.15 | 0.10 | 14.03 |
| rs4135275_rs7206012 | 6.98E-01 | 0.28 | 0.10 | 5.52E-03 | 0.04 | 0.12 | 15.91 |
| rs4135275_rs72807785 | 3.88E-01 | 0.26 | 0.09 | 5.15E-03 | -0.11 | 0.11 | 16.16 |
| rs4135275_rs77588067 | 6.86E-01 | 0.25 | 0.10 | 9.89E-03 | 0.04 | 0.11 | 13.71 |
| rs4135275_rs9922619 | 8.36E-01 | 0.15 | 0.06 | 8.89E-03 | 0.01 | 0.07 | 14.04 |
| rs4135284_rs6232 | 6.49E-01 | 0.39 | 0.17 | 1.97E-02 | 0.08 | 0.19 | 11.30 |
| rs4135284_rs78395109 | 6.12E-01 | 0.21 | 0.10 | 2.74E-02 | 0.03 | 0.11 | 10.00 |
| rs41457646_rs9922370 | 4.90E-01 | 0.32 | 0.12 | 6.30E-03 | 0.11 | 0.13 | 15.50 |
| rs4518111_rs2665271 | 1.43E-01 | 0.11 | 0.05 | 1.62E-02 | -0.07 | 0.06 | 11.83 |
| rs61781284_rs3856806 | 9.43E-02 | -0.25 | 0.10 | 1.39E-02 | 0.17 | 0.11 | 12.49 |
| rs61781284_rs7186637 | 6.64E-01 | -0.21 | 0.08 | 9.40E-03 | -0.05 | 0.10 | 13.89 |
| rs61781284_rs7190483 | 8.03E-01 | -0.27 | 0.12 | 2.62E-02 | -0.06 | 0.14 | 10.22 |
| rs62034079_rs2689264 | 2.01E-01 | 0.35 | 0.12 | 4.24E-03 | -0.14 | 0.15 | 16.99 |
| rs62034079_rs7206012 | 4.00E-01 | 0.43 | 0.16 | 8.09E-03 | 0.25 | 0.18 | 14.64 |
| rs62034079_rs967515 | 9.84E-01 | 0.38 | 0.15 | 9.31E-03 | -0.01 | 0.17 | 14.04 |
| rs6232_rs62048372 | 3.26E-01 | -0.32 | 0.13 | 1.47E-02 | -0.19 | 0.16 | 12.33 |

| | | | | | | | |
|---|---|---|---|---|---|---|---|
| rs7185938_rs1588413 | 4.95E-01 | 0.17 | 0.06 | 2.22E-03 | 0.06 | 0.07 | 19.16 |
| rs7190053_rs2665271 | 6.58E-01 | 0.13 | 0.05 | 1.99E-02 | -0.03 | 0.06 | 11.09 |
| rs7190053_rs57957459 | 9.82E-01 | 0.19 | 0.08 | 1.99E-02 | -0.02 | 0.09 | 11.12 |
| rs7190053_rs62034079 | 1.65E-01 | 0.24 | 0.10 | 1.42E-02 | -0.18 | 0.12 | 12.36 |
| rs7190483_rs71392011 | 9.87E-01 | -0.75 | 0.26 | 3.77E-03 | 0.06 | 0.28 | 18.48 |
| rs7200972_rs7194243 | 4.03E-01 | -0.21 | 0.07 | 3.03E-03 | 0.04 | 0.08 | 17.98 |
| rs7206456_rs7194243 | 1.20E-01 | -0.14 | 0.06 | 1.79E-02 | -0.11 | 0.07 | 11.49 |
| rs72921463_rs2120825 | 8.59E-01 | -0.31 | 0.13 | 1.37E-02 | -0.02 | 0.15 | 12.61 |
| rs75222427_rs9922370 | 2.18E-01 | 0.39 | 0.17 | 2.13E-02 | 0.28 | 0.18 | 11.10 |
| rs76442450_rs7190483 | 8.12E-01 | -0.45 | 0.21 | 2.84E-02 | 0.04 | 0.23 | 10.16 |
| rs77140256_rs1344503 | 3.21E-01 | -0.23 | 0.10 | 2.04E-02 | 0.07 | 0.11 | 11.07 |
| rs77140256_rs6499658 | 9.29E-01 | -0.19 | 0.08 | 1.72E-02 | 0.01 | 0.10 | 11.67 |
| rs7795794_rs17820328 | 2.20E-01 | -0.68 | 0.25 | 7.37E-03 | -0.32 | 0.31 | 15.49 |
| rs7795794_rs62048372 | 5.94E-02 | -0.29 | 0.12 | 1.80E-02 | -0.28 | 0.15 | 11.58 |
| rs9436297_rs2003583 | 1.63E-01 | 0.20 | 0.09 | 2.52E-02 | -0.12 | 0.11 | 10.28 |
| rs9436297_rs4135275 | 7.69E-01 | 0.30 | 0.09 | 1.65E-03 | 0.05 | 0.11 | 20.52 |
| rs9436297_rs41457646 | 3.28E-01 | 0.26 | 0.11 | 1.52E-02 | 0.14 | 0.12 | 12.14 |
| rs9436297_rs72807785 | 7.80E-01 | 0.27 | 0.12 | 1.82E-02 | -0.01 | 0.14 | 11.52 |
| rs9436302_rs61471078 | 2.01E-01 | -0.14 | 0.06 | 2.13E-02 | -0.09 | 0.07 | 10.85 |
| rs9436302_rs6230 | 8.82E-02 | -0.20 | 0.09 | 2.22E-02 | -0.17 | 0.11 | 10.75 |

Table 3s.4. HRS black females

| SNP | pPhenoSNP | beta_1 | SE_1 | pExpSNP | beta_2 | SE_2 | F |
|---|---|---|---|---|---|---|---|
| rs1063538_rs12918495 | 6.40E-01 | 0.43 | 0.20 | 2.90E-02 | -0.05 | 0.21 | 15.22 |
| rs1063538_rs8048396 | 4.13E-01 | 0.49 | 0.20 | 1.41E-02 | -0.11 | 0.21 | 19.44 |
| rs1076471_rs16952987 | 2.93E-01 | 0.63 | 0.29 | 2.76E-02 | -0.22 | 0.29 | 15.65 |
| rs1151999_rs12325409 | 4.23E-01 | 0.54 | 0.25 | 2.86E-02 | -0.12 | 0.27 | 15.38 |
| rs1151999_rs1421090 | 9.49E-02 | 0.59 | 0.26 | 2.03E-02 | -0.35 | 0.28 | 17.33 |
| rs1152001_rs11760956 | 5.81E-01 | 0.90 | 0.35 | 1.10E-02 | -0.05 | 0.35 | 22.28 |
| rs1152001_rs16952756 | 4.23E-01 | 0.79 | 0.37 | 3.43E-02 | 0.47 | 0.37 | 14.46 |
| rs1152001_rs708278 | 6.24E-01 | 0.73 | 0.26 | 5.50E-03 | 0.23 | 0.27 | 25.57 |
| rs1152001_rs7186521 | 8.65E-01 | 0.52 | 0.25 | 4.08E-02 | -0.02 | 0.27 | 13.30 |
| rs1152001_rs7206012 | 6.74E-01 | 0.70 | 0.26 | 8.21E-03 | 0.29 | 0.26 | 23.13 |
| rs1175542_rs6499662 | 1.02E-01 | 0.88 | 0.30 | 3.04E-03 | -0.36 | 0.32 | 30.85 |
| rs1175542_rs7206456 | 7.59E-01 | 0.86 | 0.32 | 6.84E-03 | 0.17 | 0.31 | 24.98 |

| | | | | | | | |
|---|---|---|---|---|---|---|---|
| rs1175544_rs12935710 | 3.59E-01 | 0.86 | 0.28 | 1.68E-03 | 0.31 | 0.27 | 33.67 |
| rs11760956_rs3928987 | 1.59E-01 | 0.51 | 0.24 | 3.35E-02 | -0.24 | 0.26 | 14.42 |
| rs1177809_rs1966435 | 9.23E-01 | 0.92 | 0.34 | 6.30E-03 | 0.22 | 0.33 | 25.70 |
| rs12038998_rs9921255 | 8.36E-01 | -0.64 | 0.29 | 2.65E-02 | -0.02 | 0.28 | 16.86 |
| rs12145690_rs3790427 | 2.75E-01 | -0.69 | 0.27 | 1.15E-02 | 0.19 | 0.25 | 22.19 |
| rs12145690_rs4135284 | 2.53E-01 | -0.65 | 0.28 | 1.79E-02 | 0.28 | 0.25 | 19.78 |
| rs12145690_rs4135336 | 7.29E-02 | -0.93 | 0.37 | 1.13E-02 | 0.46 | 0.31 | 24.02 |
| rs12325409_rs6499662 | 3.66E-01 | 0.56 | 0.28 | 4.44E-02 | -0.26 | 0.31 | 13.08 |
| rs12405556_rs7199363 | 9.11E-01 | -0.59 | 0.27 | 2.65E-02 | -0.07 | 0.26 | 16.77 |
| rs12447422_rs7190483 | 5.67E-01 | -0.60 | 0.26 | 2.27E-02 | 0.04 | 0.25 | 17.78 |
| rs12447422_rs9921255 | 3.97E-01 | -0.62 | 0.28 | 2.82E-02 | -0.32 | 0.29 | 16.99 |
| rs12447422_rs9924877 | 1.07E-01 | -0.75 | 0.35 | 3.34E-02 | -0.76 | 0.40 | 16.48 |
| rs12448529_rs7199363 | 8.30E-01 | -0.49 | 0.23 | 3.68E-02 | 0.03 | 0.23 | 14.45 |
| rs13428113_rs1008400 | 4.16E-01 | 0.39 | 0.18 | 3.22E-02 | -0.09 | 0.19 | 14.60 |
| rs1421090_rs16952686 | 3.92E-01 | 0.95 | 0.35 | 5.97E-03 | -0.17 | 0.36 | 25.41 |
| rs1421090_rs1966435 | 1.67E-01 | 0.57 | 0.28 | 4.19E-02 | -0.34 | 0.31 | 13.22 |
| rs1421090_rs708278 | 2.88E-01 | 0.52 | 0.22 | 1.67E-02 | -0.16 | 0.23 | 18.44 |
| rs1501299_rs7184874 | 9.00E-02 | 0.55 | 0.20 | 7.06E-03 | -0.34 | 0.23 | 23.65 |
| rs16952517_rs9921255 | 9.90E-01 | -0.85 | 0.39 | 3.10E-02 | -0.17 | 0.36 | 17.37 |
| rs16952570_rs8053279 | 2.50E-01 | -0.57 | 0.24 | 1.62E-02 | -0.33 | 0.24 | 19.23 |
| rs16952756_rs7206012 | 8.36E-01 | 0.57 | 0.27 | 3.28E-02 | 0.10 | 0.28 | 14.64 |
| rs17036321_rs7188378 | 3.41E-01 | 0.56 | 0.26 | 2.90E-02 | -0.22 | 0.28 | 15.22 |
| rs17097193_rs3811951 | 6.20E-01 | 0.82 | 0.31 | 7.67E-03 | 0.29 | 0.30 | 24.18 |
| rs17127677_rs1063538 | 4.02E-01 | 0.52 | 0.20 | 8.78E-03 | -0.15 | 0.21 | 22.32 |
| rs17127677_rs1152001 | 6.06E-01 | 1.08 | 0.37 | 3.55E-03 | -0.05 | 0.36 | 30.24 |
| rs17127677_rs1421090 | 1.77E-01 | 0.72 | 0.27 | 7.21E-03 | -0.32 | 0.29 | 23.94 |
| rs17127677_rs1805134 | 7.04E-01 | 0.60 | 0.26 | 2.20E-02 | 0.09 | 0.27 | 16.91 |
| rs17127677_rs2167270 | 4.02E-01 | 0.54 | 0.21 | 1.11E-02 | -0.16 | 0.23 | 20.87 |
| rs17127677_rs2882298 | 3.74E-01 | 0.51 | 0.22 | 2.05E-02 | 0.26 | 0.23 | 17.24 |
| rs17127677_rs4567312 | 6.61E-01 | 0.73 | 0.35 | 3.89E-02 | 0.07 | 0.36 | 14.13 |
| rs17127677_rs7206012 | 7.14E-01 | 0.59 | 0.25 | 1.71E-02 | -0.03 | 0.26 | 18.41 |
| rs17127677_rs9806929 | 7.17E-01 | 0.90 | 0.35 | 1.10E-02 | -0.05 | 0.35 | 22.28 |
| rs17127677_rs9933107 | 6.57E-01 | 0.59 | 0.24 | 1.43E-02 | -0.15 | 0.26 | 19.46 |
| rs17127677_rs9934504 | 1.55E-01 | 0.60 | 0.27 | 2.67E-02 | 0.40 | 0.27 | 15.88 |
| rs17222465_rs708262 | 8.30E-01 | -0.81 | 0.29 | 4.43E-03 | -0.10 | 0.26 | 30.10 |
| rs1805094_rs9921255 | 5.50E-01 | -1.08 | 0.40 | 6.75E-03 | -0.30 | 0.37 | 27.36 |
| rs1805094_rs9924877 | 2.44E-01 | -1.58 | 0.64 | 1.37E-02 | -0.75 | 0.58 | 25.87 |

| | | | | | | | |
|---|---|---|---|---|---|---|---|
| rs1805096_rs708262 | 2.20E-01 | -0.41 | 0.20 | 3.58E-02 | 0.11 | 0.20 | 14.31 |
| rs1861356_rs7199363 | 6.33E-01 | -0.90 | 0.30 | 2.47E-03 | -0.23 | 0.31 | 30.29 |
| rs1887285_rs11863548 | 3.87E-01 | 0.69 | 0.34 | 4.29E-02 | -0.13 | 0.36 | 13.28 |
| rs1887285_rs7193851 | 2.48E-01 | 0.78 | 0.31 | 1.24E-02 | -0.31 | 0.34 | 21.10 |
| rs2028759_rs7184874 | 4.44E-01 | 0.63 | 0.22 | 4.20E-03 | -0.10 | 0.23 | 27.23 |
| rs2028759_rs7204611 | 7.57E-01 | 0.43 | 0.20 | 3.60E-02 | 0.16 | 0.21 | 13.95 |
| rs2028760_rs1421090 | 2.62E-01 | 0.53 | 0.25 | 3.35E-02 | -0.26 | 0.28 | 14.48 |
| rs2042032_rs1344502 | 7.82E-01 | 0.62 | 0.30 | 3.76E-02 | -0.08 | 0.32 | 13.92 |
| rs2160481_rs708262 | 9.93E-01 | -0.59 | 0.28 | 3.53E-02 | -0.07 | 0.27 | 15.50 |
| rs2167270_rs1123817 | 6.08E-01 | 0.65 | 0.29 | 2.71E-02 | -0.07 | 0.31 | 15.71 |
| rs2167270_rs1966435 | 8.18E-01 | 0.69 | 0.23 | 2.82E-03 | 0.07 | 0.24 | 29.76 |
| rs2167270_rs741300 | 5.36E-01 | 0.38 | 0.19 | 4.68E-02 | -0.08 | 0.20 | 12.47 |
| rs2767485_rs7184573 | 9.84E-01 | 0.58 | 0.26 | 2.81E-02 | 0.01 | 0.28 | 15.51 |
| rs2767485_rs7184874 | 8.29E-01 | 0.62 | 0.23 | 8.46E-03 | -0.08 | 0.25 | 22.77 |
| rs2767485_rs9817428 | 6.16E-01 | 0.45 | 0.22 | 4.13E-02 | 0.14 | 0.23 | 13.21 |
| rs2882298_rs16952756 | 4.37E-01 | 0.51 | 0.22 | 2.30E-02 | 0.30 | 0.23 | 16.60 |
| rs2938395_rs12325409 | 6.94E-01 | 0.54 | 0.27 | 4.45E-02 | 0.17 | 0.27 | 12.95 |
| rs2938395_rs708278 | 6.80E-01 | 0.48 | 0.23 | 3.19E-02 | 0.09 | 0.23 | 14.71 |
| rs36072366_rs2042032 | 4.08E-01 | 0.49 | 0.25 | 4.61E-02 | 0.27 | 0.25 | 12.66 |
| rs3751813_rs1008400 | 2.01E-01 | -0.37 | 0.19 | 4.91E-02 | 0.12 | 0.19 | 12.33 |
| rs3751813_rs16952570 | 7.08E-01 | -0.58 | 0.27 | 3.21E-02 | 0.04 | 0.26 | 15.45 |
| rs3751813_rs9922708 | 6.03E-01 | -0.71 | 0.30 | 1.89E-02 | -0.26 | 0.31 | 18.06 |
| rs3811951_rs1421090 | 3.15E-01 | 0.45 | 0.23 | 4.60E-02 | -0.13 | 0.24 | 12.62 |
| rs3811951_rs7184874 | 7.32E-01 | 0.46 | 0.21 | 2.89E-02 | 0.00 | 0.22 | 15.24 |
| rs3811951_rs9806929 | 5.30E-01 | 0.60 | 0.27 | 2.67E-02 | 0.33 | 0.27 | 15.88 |
| rs3826169_rs9934504 | 5.10E-02 | 0.58 | 0.28 | 3.46E-02 | 0.67 | 0.27 | 14.57 |
| rs3928987_rs1123817 | 2.89E-01 | 0.58 | 0.28 | 4.01E-02 | -0.19 | 0.30 | 13.55 |
| rs4135284_rs1861356 | 7.48E-01 | -0.76 | 0.35 | 3.09E-02 | -0.21 | 0.34 | 16.59 |
| rs4135284_rs2665271 | 5.97E-01 | -0.53 | 0.23 | 1.82E-02 | -0.13 | 0.23 | 18.60 |
| rs4468199_rs16952756 | 7.11E-01 | 0.87 | 0.35 | 1.33E-02 | -0.09 | 0.36 | 20.68 |
| rs4606347_rs7199363 | 5.33E-01 | -0.66 | 0.30 | 2.49E-02 | -0.25 | 0.30 | 17.06 |
| rs4655537_rs12325409 | 6.95E-01 | 0.46 | 0.21 | 2.62E-02 | 0.00 | 0.22 | 15.76 |
| rs4783826_rs1966435 | 6.33E-01 | 0.43 | 0.21 | 4.04E-02 | 0.20 | 0.22 | 13.31 |
| rs55844219_rs7204690 | 5.00E-01 | -0.60 | 0.30 | 4.91E-02 | 0.02 | 0.29 | 13.15 |
| rs57669574_rs3811951 | 9.98E-01 | 0.58 | 0.28 | 3.46E-02 | 0.06 | 0.28 | 14.57 |
| rs61471078_rs1861356 | 3.95E-01 | -0.53 | 0.22 | 1.88E-02 | 0.13 | 0.22 | 18.49 |
| rs61702770_rs2042032 | 1.02E-01 | 0.89 | 0.44 | 4.06E-02 | 0.81 | 0.42 | 13.88 |
| rs6499652_rs17222465 | 9.78E-01 | -0.50 | 0.25 | 4.14E-02 | -0.11 | 0.25 | 13.39 |

| | | | | | | | |
|---|---|---|---|---|---|---|---|
| rs6499652_rs17224394 | 9.19E-01 | -0.50 | 0.24 | 4.11E-02 | -0.03 | 0.25 | 13.52 |
| rs6673591_rs6794024 | 7.41E-01 | -0.49 | 0.20 | 1.29E-02 | 0.02 | 0.20 | 20.35 |
| rs6673591_rs7204690 | 2.32E-01 | -0.44 | 0.22 | 4.24E-02 | 0.18 | 0.22 | 13.48 |
| rs6678033_rs61471078 | 4.13E-01 | -0.47 | 0.19 | 1.17E-02 | 0.07 | 0.19 | 20.83 |
| rs6773957_rs9940629 | 1.79E-01 | -0.43 | 0.18 | 1.95E-02 | 0.16 | 0.19 | 17.61 |
| rs6794024_rs12447422 | 4.89E-01 | -0.56 | 0.24 | 2.13E-02 | -0.25 | 0.25 | 18.03 |
| rs6794024_rs708262 | 8.96E-01 | -0.56 | 0.24 | 1.97E-02 | -0.10 | 0.24 | 18.17 |
| rs708258_rs2665271 | 6.99E-01 | -0.58 | 0.26 | 2.40E-02 | -0.14 | 0.26 | 16.57 |
| rs709149_rs11075994 | 4.96E-01 | 0.80 | 0.36 | 2.72E-02 | -0.16 | 0.38 | 16.47 |
| rs709149_rs1152001 | 7.30E-01 | 0.64 | 0.32 | 4.50E-02 | -0.08 | 0.34 | 13.20 |
| rs709149_rs16952756 | 8.13E-01 | 0.98 | 0.39 | 1.19E-02 | 0.03 | 0.39 | 21.66 |
| rs709149_rs7184573 | 6.18E-01 | 0.95 | 0.34 | 5.61E-03 | -0.12 | 0.35 | 27.19 |
| rs709149_rs8048396 | 9.67E-01 | 0.76 | 0.31 | 1.32E-02 | -0.05 | 0.32 | 20.36 |
| rs7184874_rs16952663 | 5.89E-01 | 0.66 | 0.31 | 2.94E-02 | 0.21 | 0.31 | 15.43 |
| rs7184874_rs16952987 | 3.53E-01 | 0.79 | 0.25 | 1.57E-03 | -0.14 | 0.26 | 34.57 |
| rs7184874_rs1966435 | 6.77E-01 | 0.58 | 0.23 | 1.02E-02 | 0.18 | 0.23 | 21.66 |
| rs7184874_rs3928987 | 1.88E-01 | 0.59 | 0.21 | 5.38E-03 | -0.18 | 0.22 | 25.41 |
| rs7184874_rs4784333 | 4.39E-01 | 0.39 | 0.20 | 4.61E-02 | 0.20 | 0.20 | 12.61 |
| rs7184874_rs708278 | 9.60E-01 | 0.54 | 0.21 | 1.14E-02 | 0.02 | 0.22 | 20.72 |
| rs7184874_rs7188378 | 5.01E-01 | 0.41 | 0.20 | 3.96E-02 | -0.14 | 0.21 | 13.43 |
| rs7184874_rs9940629 | 3.81E-01 | 0.43 | 0.20 | 2.84E-02 | -0.12 | 0.21 | 15.28 |
| rs7185938_rs12444954 | 9.70E-01 | 0.44 | 0.21 | 3.63E-02 | 0.08 | 0.22 | 13.92 |
| rs7185938_rs1588413 | 5.45E-01 | 0.56 | 0.27 | 3.75E-02 | -0.10 | 0.29 | 13.79 |
| rs7186521_rs12325409 | 2.41E-01 | 0.45 | 0.21 | 3.64E-02 | -0.23 | 0.23 | 13.91 |
| rs7186521_rs7188162 | 8.40E-01 | 0.42 | 0.22 | 5.00E-02 | -0.03 | 0.23 | 12.14 |
| rs7191513_rs11646512 | 7.34E-01 | 0.44 | 0.19 | 2.03E-02 | 0.03 | 0.20 | 17.29 |
| rs7194243_rs11646512 | 6.43E-01 | -0.42 | 0.19 | 2.87E-02 | 0.01 | 0.20 | 15.42 |
| rs7195539_rs9921255 | 8.81E-01 | -0.84 | 0.42 | 4.48E-02 | -0.07 | 0.39 | 14.31 |
| rs7199363_rs11646512 | 9.41E-01 | -0.40 | 0.19 | 3.88E-02 | -0.12 | 0.20 | 13.67 |
| rs7199363_rs8053279 | 1.99E-01 | 0.39 | 0.19 | 3.85E-02 | 0.33 | 0.20 | 13.67 |
| rs7204611_rs74022304 | 3.06E-01 | 0.57 | 0.29 | 4.97E-02 | 0.33 | 0.30 | 12.22 |
| rs7205009_rs7191513 | 7.75E-01 | 0.40 | 0.19 | 4.05E-02 | 0.08 | 0.20 | 13.33 |
| rs72805641_rs1558756 | 1.24E-01 | -0.56 | 0.27 | 3.87E-02 | 0.26 | 0.26 | 14.12 |
| rs72807785_rs8049235 | 6.65E-01 | 0.50 | 0.25 | 4.82E-02 | -0.17 | 0.27 | 12.38 |
| rs75465244_rs2197423 | 3.18E-01 | 1.07 | 0.38 | 4.28E-03 | -0.33 | 0.38 | 30.45 |
| rs7620165_rs1175544 | 1.69E-01 | 0.84 | 0.35 | 1.70E-02 | 0.46 | 0.34 | 19.38 |
| rs7626560_rs7184874 | 7.31E-01 | 0.60 | 0.29 | 3.90E-02 | 0.05 | 0.30 | 13.71 |

| | | | | | | | |
|---|---|---|---|---|---|---|---|
| rs7638903_rs3928987 | 8.43E-01 | 0.48 | 0.23 | 3.82E-02 | 0.06 | 0.24 | 13.68 |
| rs7647481_rs9934504 | 8.63E-02 | 0.76 | 0.31 | 1.51E-02 | 0.65 | 0.31 | 19.68 |
| rs8048396_rs7184874 | 9.44E-01 | 0.46 | 0.22 | 3.92E-02 | 0.01 | 0.23 | 13.51 |
| rs822395_rs7184874 | 7.62E-01 | 0.44 | 0.20 | 2.89E-02 | 0.12 | 0.21 | 15.20 |
| rs822395_rs74022304 | 5.07E-01 | 0.49 | 0.24 | 4.49E-02 | 0.24 | 0.25 | 12.84 |
| rs889904_rs7184874 | 9.65E-01 | 0.38 | 0.18 | 3.64E-02 | 0.02 | 0.19 | 13.90 |
| rs9436297_rs16952987 | 8.83E-01 | 0.99 | 0.34 | 4.20E-03 | 0.15 | 0.33 | 28.77 |
| rs9436297_rs2938395 | 9.76E-01 | 1.02 | 0.35 | 3.80E-03 | 0.03 | 0.34 | 30.43 |
| rs9436297_rs3811951 | 9.76E-01 | 0.75 | 0.28 | 7.83E-03 | 0.05 | 0.28 | 23.76 |
| rs9436297_rs7184874 | 7.29E-01 | 0.85 | 0.27 | 1.86E-03 | 0.12 | 0.27 | 33.69 |
| rs9436297_rs8048396 | 5.04E-01 | 0.87 | 0.32 | 7.29E-03 | -0.18 | 0.34 | 24.70 |
| rs9436297_rs9934504 | 4.65E-01 | 0.80 | 0.32 | 1.28E-02 | 0.28 | 0.32 | 20.63 |
| rs9436300_rs11208675 | 8.03E-01 | -0.52 | 0.25 | 3.80E-02 | -0.09 | 0.25 | 14.72 |
| rs970468_rs61471078 | 4.10E-01 | -0.41 | 0.19 | 3.39E-02 | 0.10 | 0.20 | 14.54 |
| rs9817428_rs16952663 | 6.36E-01 | 0.66 | 0.31 | 2.94E-02 | 0.11 | 0.31 | 15.43 |
| rs9922619_rs55844219 | 1.69E-01 | -0.67 | 0.31 | 2.77E-02 | -0.56 | 0.33 | 17.11 |
| rs9922708_rs708262 | 7.30E-01 | -0.61 | 0.29 | 3.96E-02 | -0.22 | 0.30 | 14.31 |
| rs9934504_rs6499662 | 8.72E-01 | 0.77 | 0.34 | 2.33E-02 | 0.11 | 0.34 | 17.12 |
| rs9934528_rs6499652 | 5.70E-02 | 0.51 | 0.23 | 2.94E-02 | 0.46 | 0.24 | 15.35 |
| rs9940629_rs12445828 | 4.35E-01 | 0.44 | 0.21 | 4.13E-02 | -0.09 | 0.23 | 13.20 |

Table 3s.5. HRS black males

| SNP | pPhenoSNP | beta_1 | SE_1 | pExpSNP | beta_2 | SE_2 | F |
|---|---|---|---|---|---|---|---|
| rs10153154_rs2665275 | 9.89E-01 | -0.64 | 0.29 | 2.67E-02 | -0.02 | 0.26 | 14.92 |
| rs10153154_rs9931580 | 8.81E-01 | -0.51 | 0.25 | 4.01E-02 | -0.14 | 0.23 | 12.48 |
| rs10521303_rs1558756 | 7.57E-01 | 0.77 | 0.33 | 1.83E-02 | 0.01 | 0.32 | 15.91 |
| rs11075994_rs1008400 | 2.78E-01 | 0.47 | 0.23 | 4.09E-02 | 0.30 | 0.23 | 11.70 |
| rs11075994_rs7190483 | 4.89E-01 | 0.62 | 0.28 | 2.75E-02 | 0.26 | 0.28 | 13.92 |
| rs11076001_rs4784333 | 3.94E-01 | -0.41 | 0.21 | 4.73E-02 | 0.08 | 0.20 | 11.14 |
| rs11076001_rs7185938 | 1.83E-01 | -0.50 | 0.23 | 2.94E-02 | 0.23 | 0.22 | 13.70 |
| rs11076001_rs7188378 | 9.88E-01 | -0.53 | 0.25 | 3.50E-02 | -0.07 | 0.24 | 12.82 |
| rs11128597_rs2197423 | 7.56E-01 | -0.77 | 0.36 | 3.22E-02 | -0.19 | 0.31 | 14.71 |
| rs11128597_rs9934504 | 3.28E-01 | -0.78 | 0.35 | 2.80E-02 | 0.19 | 0.31 | 14.98 |
| rs11208654_rs7185938 | 3.30E-01 | -0.52 | 0.24 | 3.46E-02 | 0.16 | 0.23 | 13.11 |
| rs11801408_rs1861356 | 9.90E-01 | 0.50 | 0.24 | 3.59E-02 | 0.08 | 0.24 | 12.33 |

| | | | | | | | |
|---|---|---|---|---|---|---|---|
| rs12409877_rs9931580 | 2.87E-01 | -0.55 | 0.22 | 1.48E-02 | -0.31 | 0.22 | 17.23 |
| rs12495941_rs1861356 | 5.77E-01 | 0.59 | 0.24 | 1.42E-02 | -0.06 | 0.24 | 17.19 |
| rs12935710_rs1861356 | 2.33E-01 | 0.46 | 0.23 | 4.33E-02 | 0.34 | 0.23 | 11.41 |
| rs1327121_rs16861205 | 6.81E-01 | -0.64 | 0.32 | 4.15E-02 | -0.03 | 0.27 | 13.01 |
| rs1327121_rs2024470 | 4.14E-01 | -0.53 | 0.26 | 4.68E-02 | 0.12 | 0.24 | 11.73 |
| rs13428113_rs709149 | 6.77E-01 | 0.54 | 0.25 | 2.92E-02 | 0.16 | 0.24 | 13.45 |
| rs1345390_rs16953089 | 6.94E-01 | -0.58 | 0.22 | 8.25E-03 | 0.00 | 0.21 | 20.57 |
| rs1421090_rs2665275 | 2.26E-01 | -0.67 | 0.28 | 1.63E-02 | 0.25 | 0.24 | 18.03 |
| rs1421090_rs55844219 | 2.36E-01 | -0.77 | 0.31 | 1.24E-02 | 0.13 | 0.27 | 19.61 |
| rs1421090_rs8053279 | 5.59E-01 | -0.64 | 0.23 | 6.04E-03 | 0.06 | 0.22 | 22.35 |
| rs1501299_rs12447422 | 4.09E-01 | -0.67 | 0.29 | 1.87E-02 | -0.34 | 0.27 | 16.87 |
| rs1501299_rs3811951 | 1.13E-01 | -0.50 | 0.25 | 4.93E-02 | -0.40 | 0.25 | 11.27 |
| rs16952594_rs7199363 | 1.56E-01 | -0.71 | 0.33 | 3.30E-02 | -0.57 | 0.32 | 14.04 |
| rs17127673_rs1805134 | 2.45E-01 | 0.47 | 0.22 | 3.33E-02 | 0.31 | 0.22 | 12.67 |
| rs1805096_rs8053279 | 8.21E-01 | 0.56 | 0.21 | 9.42E-03 | -0.02 | 0.21 | 19.47 |
| rs1805134_rs7186220 | 5.51E-02 | 0.48 | 0.22 | 2.95E-02 | 0.46 | 0.22 | 13.28 |
| rs1861356_rs1008400 | 1.36E-01 | 0.53 | 0.24 | 3.08E-02 | 0.40 | 0.24 | 13.23 |
| rs1861356_rs7186220 | 1.15E-01 | 0.53 | 0.24 | 2.94E-02 | 0.45 | 0.24 | 13.36 |
| rs1938496_rs1125392 | 9.11E-01 | -0.94 | 0.32 | 3.82E-03 | -0.14 | 0.27 | 28.30 |
| rs1938496_rs3811951 | 5.40E-01 | -0.94 | 0.37 | 1.09E-02 | -0.25 | 0.31 | 22.28 |
| rs1938496_rs55844219 | 9.81E-01 | -1.01 | 0.41 | 1.23E-02 | -0.20 | 0.33 | 21.38 |
| rs1938496_rs6499640 | 1.60E-01 | -0.65 | 0.32 | 3.96E-02 | -0.52 | 0.31 | 12.83 |
| rs1938496_rs7188378 | 9.93E-01 | -0.65 | 0.28 | 2.01E-02 | -0.11 | 0.26 | 16.07 |
| rs1938496_rs7203572 | 3.86E-01 | -0.69 | 0.32 | 3.18E-02 | -0.38 | 0.30 | 14.36 |
| rs1938496_rs9817428 | 2.94E-01 | -0.79 | 0.32 | 1.35E-02 | -0.39 | 0.29 | 19.38 |
| rs2067819_rs1125392 | 3.71E-01 | -0.58 | 0.27 | 3.02E-02 | 0.14 | 0.24 | 14.14 |
| rs2067819_rs1421090 | 7.47E-01 | -0.78 | 0.34 | 1.96E-02 | -0.25 | 0.30 | 17.12 |
| rs2067819_rs3811951 | 9.64E-01 | -0.72 | 0.32 | 2.63E-02 | -0.06 | 0.29 | 15.29 |
| rs2067819_rs7206010 | 9.53E-01 | -0.63 | 0.29 | 3.10E-02 | -0.07 | 0.26 | 14.04 |
| rs36219760_rs7206010 | 6.01E-01 | -0.72 | 0.30 | 1.50E-02 | 0.04 | 0.26 | 18.77 |
| rs3790427_rs4784333 | 1.48E-01 | -0.47 | 0.23 | 4.00E-02 | 0.19 | 0.22 | 12.22 |
| rs4145574_rs2665275 | 9.79E-01 | -0.76 | 0.37 | 3.67E-02 | -0.06 | 0.31 | 13.83 |
| rs436321_rs11644943 | 4.76E-01 | 0.57 | 0.27 | 3.47E-02 | 0.24 | 0.27 | 12.57 |

| | | | | | | | |
|---|---|---|---|---|---|---|---|
| rs4567312_rs7201878 | 4.29E-01 | 0.64 | 0.30 | 2.95E-02 | -0.09 | 0.30 | 13.40 |
| rs4655802_rs8053279 | 2.65E-01 | 0.44 | 0.22 | 4.08E-02 | -0.24 | 0.21 | 11.78 |
| rs4655802_rs9931580 | 2.32E-01 | -0.52 | 0.25 | 3.68E-02 | 0.20 | 0.23 | 12.66 |
| rs61471078_rs8053279 | 8.52E-01 | -0.55 | 0.22 | 1.33E-02 | -0.09 | 0.21 | 17.77 |
| rs62048372_rs7184573 | 5.57E-02 | 0.73 | 0.30 | 1.36E-02 | -0.44 | 0.31 | 17.66 |
| rs6230_rs10852525 | 1.77E-01 | 0.59 | 0.28 | 3.44E-02 | 0.39 | 0.28 | 12.66 |
| rs6233_rs1861555 | 5.44E-01 | 0.56 | 0.27 | 3.37E-02 | 0.18 | 0.26 | 12.71 |
| rs6499640_rs12447422 | 7.49E-01 | -0.56 | 0.26 | 3.25E-02 | -0.16 | 0.24 | 13.74 |
| rs6499640_rs2665275 | 3.85E-01 | -0.63 | 0.27 | 1.82E-02 | 0.19 | 0.24 | 17.15 |
| rs6499652_rs7194907 | 2.17E-01 | -0.55 | 0.24 | 2.02E-02 | 0.18 | 0.23 | 15.62 |
| rs6673591_rs1861555 | 5.46E-01 | 0.52 | 0.24 | 2.72E-02 | 0.17 | 0.23 | 13.74 |
| rs6782178_rs1558756 | 8.33E-01 | 0.56 | 0.24 | 1.86E-02 | 0.05 | 0.23 | 15.79 |
| rs6782178_rs7194907 | 3.16E-01 | -0.56 | 0.22 | 1.09E-02 | -0.29 | 0.21 | 18.96 |
| rs6794024_rs55844219 | 8.52E-01 | -0.55 | 0.26 | 3.31E-02 | -0.05 | 0.23 | 13.71 |
| rs7185938_rs8053279 | 4.48E-01 | 0.49 | 0.21 | 1.83E-02 | -0.11 | 0.21 | 15.80 |
| rs7186637_rs12919488 | 1.30E-01 | 0.63 | 0.26 | 1.69E-02 | -0.30 | 0.27 | 16.39 |
| rs7186637_rs12935710 | 6.00E-01 | 0.43 | 0.21 | 4.56E-02 | 0.19 | 0.21 | 11.16 |
| rs7186637_rs1861356 | 3.05E-01 | 0.48 | 0.24 | 4.46E-02 | -0.17 | 0.24 | 11.26 |
| rs7188378_rs8053279 | 8.80E-01 | 0.45 | 0.22 | 3.72E-02 | 0.07 | 0.22 | 12.18 |
| rs7194907_rs11863548 | 1.50E-01 | 0.58 | 0.27 | 3.01E-02 | -0.30 | 0.27 | 13.29 |
| rs7206010_rs16953089 | 7.27E-01 | -0.67 | 0.29 | 1.87E-02 | 0.01 | 0.26 | 16.87 |
| rs7206010_rs9931580 | 8.31E-01 | -0.54 | 0.24 | 2.50E-02 | -0.13 | 0.23 | 14.82 |
| rs741300_rs8053279 | 7.41E-01 | -0.99 | 0.40 | 1.24E-02 | 0.04 | 0.39 | 18.08 |
| rs7602_rs12447422 | 7.59E-01 | -0.61 | 0.28 | 2.85E-02 | -0.14 | 0.26 | 14.16 |
| rs8061518_rs8053279 | 3.00E-01 | 0.46 | 0.22 | 3.64E-02 | -0.20 | 0.22 | 12.32 |
| rs9817428_rs16953089 | 7.03E-01 | -0.68 | 0.30 | 2.21E-02 | 0.02 | 0.26 | 16.36 |
| rs9817428_rs55844219 | 6.90E-01 | -0.54 | 0.27 | 4.62E-02 | -0.17 | 0.25 | 11.74 |
| rs9934504_rs72807785 | 8.59E-01 | -1.33 | 0.49 | 6.93E-03 | -0.15 | 0.36 | 28.06 |
| rs9934504_rs8053279 | 3.98E-01 | -0.66 | 0.25 | 9.01E-03 | 0.15 | 0.24 | 20.23 |
| rs9940629_rs1008400 | 2.00E-01 | 0.46 | 0.20 | 2.33E-02 | 0.26 | 0.20 | 14.61 |
| rs9940629_rs1861356 | 4.60E-01 | 0.56 | 0.24 | 1.89E-02 | 0.20 | 0.23 | 15.63 |

Table 3s.6. HRS black non-stratified

| SNP | pPhenoSNP | beta_1 | SE_1 | pExpSNP | beta_2 | SE_2 | F |
|---|---|---|---|---|---|---|---|
| rs10157915_rs1938496 | 4.44E-01 | -0.79 | 0.39 | 4.11E-02 | -0.29 | 0.35 | 14.82 |
| rs10157915_rs7204690 | 7.04E-01 | -1.11 | 0.50 | 2.63E-02 | -0.24 | 0.42 | 18.42 |
| rs1063538_rs4784330 | 7.36E-01 | 0.31 | 0.15 | 4.31E-02 | 0.07 | 0.16 | 12.37 |
| rs1063538_rs8048396 | 5.85E-01 | 0.33 | 0.14 | 2.00E-02 | -0.05 | 0.15 | 16.46 |
| rs1063538_rs9940629 | 9.50E-01 | 0.34 | 0.14 | 1.44E-02 | 0.02 | 0.14 | 18.38 |
| rs10852521_rs4784330 | 2.83E-01 | 0.55 | 0.23 | 1.61E-02 | -0.14 | 0.24 | 17.79 |
| rs10852525_rs708278 | 2.63E-01 | 0.44 | 0.18 | 1.56E-02 | 0.22 | 0.18 | 17.84 |
| rs11075994_rs1966435 | 9.67E-01 | 0.44 | 0.21 | 3.77E-02 | 0.12 | 0.22 | 13.16 |
| rs11076001_rs6567166 | 8.87E-01 | -0.53 | 0.24 | 2.94E-02 | -0.08 | 0.23 | 15.59 |
| rs11208654_rs6794024 | 8.12E-01 | -0.52 | 0.21 | 1.15E-02 | 0.01 | 0.20 | 20.75 |
| rs11208654_rs708262 | 4.11E-01 | -0.39 | 0.19 | 3.99E-02 | 0.14 | 0.18 | 13.31 |
| rs11208654_rs9308762 | 8.52E-01 | -0.78 | 0.31 | 1.27E-02 | 0.04 | 0.27 | 21.77 |
| rs1151999_rs10852525 | 9.01E-01 | 0.49 | 0.21 | 1.89E-02 | 0.03 | 0.21 | 16.91 |
| rs1151999_rs12918495 | 6.97E-01 | 0.42 | 0.18 | 2.02E-02 | 0.13 | 0.18 | 16.43 |
| rs1151999_rs7184874 | 8.49E-01 | 0.51 | 0.18 | 3.52E-03 | -0.01 | 0.18 | 26.41 |
| rs1152001_rs7206012 | 1.88E-01 | 0.40 | 0.20 | 4.00E-02 | -0.14 | 0.21 | 12.76 |
| rs1175542_rs16952594 | 4.69E-01 | 0.70 | 0.29 | 1.50E-02 | 0.24 | 0.29 | 18.26 |
| rs1175544_rs11075994 | 8.05E-01 | 0.89 | 0.33 | 7.66E-03 | -0.03 | 0.34 | 22.45 |
| rs1175544_rs436321 | 9.35E-01 | 0.44 | 0.21 | 3.24E-02 | 0.00 | 0.21 | 13.92 |
| rs1175544_rs7193851 | 3.75E-01 | 0.59 | 0.22 | 8.27E-03 | 0.26 | 0.22 | 21.61 |
| rs1177809_rs8048396 | 7.11E-01 | 0.62 | 0.23 | 6.27E-03 | 0.07 | 0.23 | 23.15 |
| rs12145690_rs1938496 | 4.05E-01 | -0.50 | 0.21 | 1.71E-02 | 0.09 | 0.20 | 18.31 |
| rs12145690_rs2160481 | 4.86E-01 | -0.61 | 0.21 | 3.40E-03 | -0.20 | 0.20 | 28.70 |
| rs12145690_rs4655802 | 3.90E-01 | -0.48 | 0.22 | 2.88E-02 | -0.24 | 0.22 | 14.44 |
| rs12145690_rs708262 | 8.65E-02 | -0.34 | 0.15 | 2.29E-02 | 0.20 | 0.15 | 16.09 |
| rs12145690_rs7204690 | 3.16E-01 | -0.54 | 0.20 | 7.95E-03 | 0.13 | 0.19 | 22.75 |
| rs12447422_rs708262 | 9.19E-02 | -0.32 | 0.16 | 4.90E-02 | 0.20 | 0.16 | 12.02 |
| rs12447422_rs7206456 | 5.85E-01 | -0.54 | 0.25 | 2.94E-02 | 0.04 | 0.23 | 15.48 |
| rs12447422_rs9921255 | 8.86E-01 | -0.58 | 0.22 | 8.49E-03 | -0.12 | 0.21 | 23.12 |
| rs12447422_rs9924877 | 1.35E-01 | -0.63 | 0.26 | 1.67E-02 | -0.53 | 0.27 | 19.31 |
| rs12935710_rs1588413 | 9.14E-02 | 0.58 | 0.22 | 7.51E-03 | 0.41 | 0.22 | 22.00 |
| rs12935710_rs7194907 | 5.33E-01 | 0.32 | 0.14 | 2.45E-02 | 0.11 | 0.15 | 15.40 |

| | | | | | | | |
|---|---|---|---|---|---|---|---|
| rs12935710_rs7201878 | 4.04E-01 | 0.34 | 0.17 | 4.27E-02 | -0.08 | 0.17 | 12.47 |
| rs13061415_rs1861555 | 7.60E-01 | 0.50 | 0.20 | 1.41E-02 | 0.08 | 0.21 | 18.49 |
| rs13099634_rs9928094 | 6.89E-01 | -0.42 | 0.21 | 4.52E-02 | -0.18 | 0.21 | 12.58 |
| rs13169290_rs2160481 | 8.00E-01 | -0.79 | 0.33 | 1.78E-02 | 0.00 | 0.28 | 19.95 |
| rs1344502_rs12325409 | 7.62E-01 | 0.35 | 0.17 | 4.18E-02 | -0.06 | 0.18 | 12.50 |
| rs1344502_rs1588413 | 3.02E-01 | 0.64 | 0.26 | 1.60E-02 | -0.24 | 0.28 | 18.12 |
| rs1344502_rs77345086 | 6.79E-01 | 0.45 | 0.23 | 4.76E-02 | -0.04 | 0.24 | 11.90 |
| rs1421090_rs12447422 | 6.97E-01 | -0.50 | 0.20 | 1.53E-02 | -0.03 | 0.20 | 18.92 |
| rs16861205_rs3828942 | 4.62E-01 | -0.63 | 0.25 | 1.16E-02 | 0.02 | 0.23 | 21.16 |
| rs16952517_rs2160481 | 2.45E-01 | -0.78 | 0.31 | 1.20E-02 | -0.47 | 0.30 | 22.80 |
| rs16952686_rs7194907 | 8.96E-01 | 0.39 | 0.17 | 1.98E-02 | 0.05 | 0.17 | 16.50 |
| rs17036321_rs1175542 | 1.69E-01 | 0.56 | 0.21 | 8.42E-03 | -0.27 | 0.23 | 21.57 |
| rs17036321_rs2003583 | 2.80E-01 | 0.61 | 0.25 | 1.68E-02 | -0.24 | 0.27 | 17.53 |
| rs17036321_rs2882298 | 8.83E-01 | 0.43 | 0.20 | 2.93E-02 | 0.11 | 0.20 | 14.41 |
| rs17036321_rs4783826 | 2.07E-01 | 0.44 | 0.20 | 3.05E-02 | -0.22 | 0.22 | 14.20 |
| rs17036321_rs9933107 | 3.83E-01 | 0.76 | 0.24 | 1.64E-03 | -0.10 | 0.25 | 31.09 |
| rs17127677_rs1063538 | 9.34E-01 | 0.38 | 0.15 | 1.27E-02 | 0.04 | 0.16 | 18.97 |
| rs17127677_rs1421090 | 9.74E-01 | 0.43 | 0.19 | 2.41E-02 | 0.03 | 0.20 | 15.45 |
| rs17127677_rs1805134 | 1.26E-01 | 0.57 | 0.19 | 2.66E-03 | 0.33 | 0.19 | 28.06 |
| rs17127677_rs2960422 | 3.26E-01 | 0.38 | 0.17 | 2.34E-02 | 0.20 | 0.17 | 15.59 |
| rs17127677_rs4567312 | 2.56E-01 | 0.60 | 0.27 | 2.43E-02 | 0.28 | 0.27 | 15.74 |
| rs17127677_rs7184573 | 7.23E-01 | 0.71 | 0.21 | 7.65E-04 | -0.03 | 0.21 | 36.43 |
| rs17127677_rs7186521 | 2.19E-01 | 0.40 | 0.16 | 1.62E-02 | 0.21 | 0.17 | 17.59 |
| rs17222465_rs708262 | 9.75E-01 | -0.41 | 0.19 | 3.40E-02 | -0.07 | 0.19 | 14.17 |
| rs17226942_rs2075202 | 1.32E-01 | -0.62 | 0.29 | 2.99E-02 | 0.31 | 0.25 | 15.80 |
| rs17817469_rs9922370 | 1.31E-01 | -0.52 | 0.25 | 4.13E-02 | -0.47 | 0.27 | 13.47 |
| rs1805134_rs60386982 | 7.37E-01 | 0.49 | 0.20 | 1.53E-02 | 0.09 | 0.21 | 18.00 |
| rs1861555_rs11863548 | 2.62E-01 | 0.50 | 0.23 | 3.39E-02 | -0.16 | 0.25 | 13.75 |
| rs1861555_rs16952987 | 9.98E-01 | 0.58 | 0.22 | 9.28E-03 | 0.03 | 0.23 | 21.16 |
| rs1861555_rs77345086 | 5.48E-01 | 0.83 | 0.36 | 2.04E-02 | 0.36 | 0.35 | 16.83 |
| rs1861867_rs55844219 | 5.87E-01 | -0.46 | 0.22 | 3.55E-02 | 0.04 | 0.21 | 14.27 |
| rs1861868_rs16953039 | 9.10E-01 | 0.61 | 0.27 | 2.58E-02 | 0.04 | 0.28 | 15.43 |
| rs1938496_rs1559509 | 8.65E-01 | -0.47 | 0.23 | 3.65E-02 | -0.08 | 0.22 | 14.06 |

| | | | | | | | |
|---|---|---|---|---|---|---|---|
| rs1938496_rs16861205 | 4.98E-01 | -0.52 | 0.26 | 4.69E-02 | 0.08 | 0.24 | 12.95 |
| rs1938496_rs16952730 | 6.12E-01 | -0.50 | 0.24 | 3.56E-02 | 0.02 | 0.22 | 14.23 |
| rs1938496_rs7190492 | 7.74E-01 | -0.72 | 0.34 | 3.63E-02 | -0.06 | 0.31 | 14.99 |
| rs1938496_rs7204690 | 7.78E-01 | -0.88 | 0.33 | 7.78E-03 | -0.11 | 0.28 | 26.06 |
| rs2042032_rs12444954 | 2.07E-01 | 0.49 | 0.24 | 4.20E-02 | 0.41 | 0.24 | 12.55 |
| rs2160481_rs12931859 | 7.79E-01 | -0.70 | 0.36 | 4.94E-02 | -0.15 | 0.32 | 13.58 |
| rs2160481_rs16953089 | 4.86E-01 | -0.88 | 0.29 | 2.63E-03 | -0.33 | 0.27 | 32.72 |
| rs2160481_rs17227068 | 3.05E-01 | -0.99 | 0.40 | 1.28E-02 | 0.21 | 0.32 | 22.78 |
| rs2160481_rs2665275 | 4.06E-01 | -0.44 | 0.22 | 4.81E-02 | -0.27 | 0.22 | 12.63 |
| rs2160481_rs34114122 | 4.40E-01 | -0.79 | 0.39 | 4.36E-02 | -0.38 | 0.38 | 13.73 |
| rs2167270_rs10521307 | 7.86E-02 | 0.34 | 0.16 | 3.05E-02 | -0.23 | 0.16 | 14.15 |
| rs2167270_rs1588413 | 6.48E-01 | 0.57 | 0.21 | 6.57E-03 | -0.03 | 0.22 | 23.04 |
| rs2167270_rs1966435 | 1.74E-01 | 0.46 | 0.17 | 6.41E-03 | -0.17 | 0.18 | 22.83 |
| rs2167270_rs7194907 | 1.54E-01 | 0.32 | 0.14 | 2.16E-02 | -0.19 | 0.15 | 16.07 |
| rs2665271_rs8053279 | 9.47E-01 | 0.33 | 0.14 | 1.64E-02 | 0.02 | 0.14 | 17.53 |
| rs271923_rs12448529 | 4.57E-01 | -0.91 | 0.38 | 1.79E-02 | -0.42 | 0.35 | 20.86 |
| rs2767485_rs10852525 | 1.78E-01 | 0.44 | 0.20 | 3.01E-02 | 0.29 | 0.20 | 14.27 |
| rs2882298_rs1344502 | 8.40E-01 | 0.36 | 0.16 | 2.15E-02 | 0.08 | 0.16 | 16.03 |
| rs2938394_rs3826169 | 2.24E-01 | 0.80 | 0.32 | 1.11E-02 | 0.39 | 0.31 | 20.52 |
| rs2938394_rs7185735 | 4.56E-01 | 0.39 | 0.17 | 2.07E-02 | -0.12 | 0.17 | 16.24 |
| rs36072366_rs9940629 | 4.15E-01 | 0.37 | 0.14 | 9.12E-03 | 0.19 | 0.15 | 20.85 |
| rs3790427_rs16952517 | 7.67E-01 | -0.50 | 0.24 | 4.03E-02 | -0.02 | 0.22 | 13.88 |
| rs3790427_rs16952649 | 2.01E-01 | -0.52 | 0.22 | 1.76E-02 | 0.14 | 0.20 | 18.47 |
| rs3790427_rs4784333 | 9.73E-02 | -0.34 | 0.16 | 2.70E-02 | 0.20 | 0.15 | 15.15 |
| rs3811951_rs73617811 | 5.72E-01 | 0.51 | 0.23 | 3.07E-02 | -0.01 | 0.24 | 14.23 |
| rs3826169_rs1861555 | 5.91E-01 | 0.98 | 0.36 | 6.94E-03 | 0.26 | 0.35 | 23.98 |
| rs3826169_rs55844219 | 7.98E-02 | -0.61 | 0.29 | 3.58E-02 | 0.31 | 0.27 | 14.09 |
| rs3828942_rs16952517 | 8.09E-01 | -0.90 | 0.31 | 3.82E-03 | -0.07 | 0.27 | 29.73 |
| rs3828942_rs2160481 | 6.09E-01 | -0.86 | 0.31 | 5.36E-03 | -0.24 | 0.27 | 28.92 |
| rs3928987_rs1588413 | 1.09E-01 | 0.72 | 0.26 | 6.30E-03 | -0.26 | 0.28 | 22.77 |
| rs4135294_rs2665275 | 4.37E-01 | -0.74 | 0.28 | 8.87E-03 | -0.30 | 0.26 | 24.21 |
| rs4135294_rs9921255 | 6.70E-01 | -0.94 | 0.34 | 5.73E-03 | -0.20 | 0.29 | 27.90 |
| rs4135361_rs1152001 | 6.62E-01 | 0.79 | 0.29 | 6.05E-03 | 0.02 | 0.28 | 24.68 |

| | | | | | | | |
|---|---|---|---|---|---|---|---|
| rs4135361_rs7191718 | 9.71E-01 | 0.38 | 0.19 | 4.59E-02 | 0.10 | 0.19 | 12.03 |
| rs4468199_rs11863548 | 7.02E-01 | 0.60 | 0.24 | 1.08E-02 | 0.03 | 0.24 | 20.16 |
| rs4567312_rs11075994 | 6.95E-01 | 0.78 | 0.30 | 1.03E-02 | 0.15 | 0.29 | 21.22 |
| rs4567312_rs61702770 | 6.66E-01 | 0.55 | 0.27 | 4.51E-02 | 0.18 | 0.27 | 12.41 |
| rs4567312_rs75518380 | 9.40E-01 | 0.87 | 0.37 | 1.72E-02 | 0.11 | 0.36 | 18.26 |
| rs4655537_rs7205986 | 9.92E-01 | 0.41 | 0.16 | 1.14E-02 | 0.04 | 0.17 | 19.53 |
| rs4784323_rs10852525 | 2.26E-01 | 0.55 | 0.28 | 4.70E-02 | -0.24 | 0.30 | 12.10 |
| rs4784323_rs9925952 | 1.28E-01 | 0.79 | 0.34 | 1.89E-02 | 0.62 | 0.33 | 17.13 |
| rs4784330_rs8049235 | 2.94E-01 | 0.37 | 0.16 | 1.69E-02 | 0.16 | 0.16 | 17.37 |
| rs60386982_rs3826169 | 3.03E-01 | 1.08 | 0.35 | 2.26E-03 | 0.44 | 0.32 | 31.97 |
| rs61702770_rs2042032 | 7.92E-02 | 0.61 | 0.28 | 3.00E-02 | 0.63 | 0.28 | 14.53 |
| rs6233_rs79977114 | 4.10E-01 | 0.44 | 0.21 | 3.29E-02 | 0.28 | 0.21 | 13.80 |
| rs6233_rs9940629 | 8.48E-01 | 0.40 | 0.14 | 3.95E-03 | 0.08 | 0.14 | 25.59 |
| rs6669354_rs11075994 | 3.38E-01 | 0.69 | 0.32 | 2.97E-02 | 0.38 | 0.32 | 14.33 |
| rs6669354_rs12325409 | 3.77E-01 | 0.49 | 0.23 | 3.53E-02 | 0.26 | 0.23 | 13.49 |
| rs6673591_rs72807785 | 3.33E-01 | -0.37 | 0.19 | 4.85E-02 | 0.12 | 0.18 | 12.13 |
| rs6768587_rs2665275 | 3.04E-01 | -0.41 | 0.19 | 2.86E-02 | -0.26 | 0.19 | 15.14 |
| rs6782178_rs1152001 | 9.95E-01 | 0.42 | 0.16 | 9.21E-03 | 0.06 | 0.16 | 20.76 |
| rs6782178_rs8048396 | 3.90E-01 | 0.28 | 0.14 | 4.37E-02 | 0.12 | 0.14 | 12.30 |
| rs6794024_rs28410925 | 6.47E-01 | -0.63 | 0.26 | 1.63E-02 | -0.01 | 0.24 | 19.22 |
| rs708258_rs2665271 | 6.10E-01 | -0.39 | 0.18 | 3.19E-02 | -0.14 | 0.19 | 14.12 |
| rs709149_rs11646488 | 4.44E-01 | 0.82 | 0.34 | 1.54E-02 | -0.15 | 0.35 | 18.57 |
| rs709149_rs1558756 | 7.88E-01 | 0.67 | 0.28 | 1.62E-02 | 0.09 | 0.28 | 17.87 |
| rs709149_rs16952756 | 5.76E-01 | 0.65 | 0.29 | 2.19E-02 | 0.19 | 0.29 | 16.31 |
| rs709149_rs16952987 | 5.79E-01 | 0.58 | 0.24 | 1.59E-02 | -0.11 | 0.25 | 17.94 |
| rs709149_rs189554127 | 9.03E-01 | 0.65 | 0.30 | 3.06E-02 | -0.07 | 0.31 | 14.23 |
| rs709149_rs708258 | 6.51E-01 | 0.46 | 0.23 | 4.14E-02 | -0.15 | 0.24 | 12.58 |
| rs709149_rs7184573 | 2.06E-01 | 0.63 | 0.25 | 1.14E-02 | -0.31 | 0.27 | 20.03 |
| rs709149_rs8044769 | 8.02E-01 | 0.59 | 0.26 | 2.28E-02 | 0.05 | 0.26 | 15.94 |
| rs7184874_rs10852521 | 1.12E-01 | 0.49 | 0.18 | 7.36E-03 | 0.32 | 0.18 | 22.10 |
| rs7184874_rs1588413 | 4.17E-01 | 0.65 | 0.24 | 6.91E-03 | 0.21 | 0.24 | 23.08 |
| rs7184874_rs16952634 | 2.86E-01 | 0.51 | 0.21 | 1.34E-02 | 0.27 | 0.21 | 18.76 |
| rs7184874_rs3928987 | 4.08E-01 | 0.33 | 0.16 | 3.19E-02 | -0.08 | 0.16 | 13.92 |

| | | | | | | | |
|---|---|---|---|---|---|---|---|
| rs7184874_rs9940629 | 6.86E-01 | 0.35 | 0.15 | 1.76E-02 | 0.07 | 0.15 | 17.12 |
| rs7185735_rs9940629 | 3.34E-01 | 0.41 | 0.14 | 3.48E-03 | -0.09 | 0.14 | 26.39 |
| rs7191718_rs9940629 | 5.30E-01 | 0.26 | 0.13 | 4.84E-02 | 0.14 | 0.14 | 11.82 |
| rs7195539_rs12447422 | 9.07E-01 | -0.59 | 0.28 | 3.29E-02 | -0.13 | 0.26 | 15.03 |
| rs7199363_rs16953065 | 9.09E-02 | -0.59 | 0.27 | 2.83E-02 | -0.54 | 0.28 | 15.45 |
| rs7200972_rs7199363 | 5.31E-01 | -0.63 | 0.26 | 1.61E-02 | -0.25 | 0.26 | 18.32 |
| rs7206010_rs11646512 | 5.24E-01 | -0.30 | 0.14 | 3.33E-02 | 0.01 | 0.15 | 13.84 |
| rs7206010_rs2665275 | 3.57E-01 | -0.41 | 0.17 | 1.72E-02 | 0.07 | 0.17 | 17.87 |
| rs7206790_rs2665275 | 6.42E-01 | -0.29 | 0.15 | 4.63E-02 | -0.02 | 0.15 | 12.14 |
| rs72803661_rs2160481 | 1.06E-01 | -0.91 | 0.38 | 1.62E-02 | -0.66 | 0.38 | 20.69 |
| rs72921463_rs9302654 | 6.48E-01 | 0.64 | 0.31 | 4.35E-02 | -0.10 | 0.33 | 12.50 |
| rs72925327_rs1177809 | 5.18E-01 | 0.86 | 0.32 | 7.14E-03 | 0.24 | 0.32 | 22.71 |
| rs72925327_rs1861555 | 5.45E-01 | 1.21 | 0.35 | 4.68E-04 | 0.35 | 0.30 | 44.18 |
| rs72925327_rs2042032 | 2.30E-01 | 0.74 | 0.34 | 2.92E-02 | 0.44 | 0.34 | 14.69 |
| rs73609956_rs1344503 | 8.44E-01 | -0.87 | 0.38 | 2.10E-02 | -0.14 | 0.33 | 18.91 |
| rs73609956_rs6499652 | 3.50E-01 | -0.37 | 0.18 | 4.22E-02 | 0.09 | 0.18 | 12.76 |
| rs74022304_rs9940629 | 9.91E-01 | 0.50 | 0.22 | 1.94E-02 | 0.02 | 0.22 | 16.61 |
| rs74081930_rs1966435 | 6.52E-01 | 0.72 | 0.29 | 1.38E-02 | -0.03 | 0.30 | 19.27 |
| rs74081930_rs8048396 | 9.03E-01 | 0.51 | 0.25 | 4.27E-02 | 0.07 | 0.26 | 12.50 |
| rs7555955_rs11644943 | 2.07E-01 | 0.50 | 0.25 | 4.19E-02 | 0.36 | 0.24 | 12.72 |
| rs76442450_rs2160481 | 7.30E-01 | -0.65 | 0.27 | 1.74E-02 | -0.21 | 0.26 | 19.23 |
| rs7647481_rs10852525 | 6.75E-01 | 0.54 | 0.23 | 2.01E-02 | 0.13 | 0.23 | 16.75 |
| rs78961312_rs9871668 | 8.81E-01 | -1.53 | 0.50 | 2.07E-03 | -0.17 | 0.38 | 38.78 |
| rs79977114_rs7186637 | 5.08E-01 | 0.41 | 0.20 | 4.50E-02 | 0.19 | 0.21 | 12.16 |
| rs79977114_rs7193851 | 9.85E-02 | 0.56 | 0.26 | 3.32E-02 | 0.56 | 0.26 | 13.78 |
| rs79977114_rs7204611 | 5.22E-01 | 0.49 | 0.22 | 2.31E-02 | 0.19 | 0.22 | 15.75 |
| rs8048396_rs10521303 | 6.21E-01 | 0.32 | 0.15 | 3.29E-02 | 0.12 | 0.15 | 13.78 |
| rs8048396_rs7185938 | 8.60E-01 | 0.35 | 0.15 | 1.83E-02 | 0.08 | 0.15 | 16.96 |
| rs8048396_rs9924983 | 9.07E-01 | 0.31 | 0.15 | 3.61E-02 | 0.01 | 0.15 | 13.29 |
| rs822396_rs11076001 | 7.29E-01 | -0.40 | 0.19 | 3.56E-02 | -0.10 | 0.19 | 14.04 |
| rs9436297_rs11863548 | 7.69E-01 | 0.47 | 0.22 | 2.96E-02 | -0.01 | 0.23 | 14.46 |
| rs9436297_rs1861555 | 8.67E-01 | 1.05 | 0.32 | 8.75E-04 | 0.09 | 0.29 | 38.40 |
| rs9436297_rs1966435 | 7.07E-01 | 0.69 | 0.25 | 6.25E-03 | -0.06 | 0.26 | 23.75 |

| | | | | | | | |
|---|---|---|---|---|---|---|---|
| rs9436297_rs60386982 | 4.67E-01 | 0.83 | 0.28 | 3.06E-03 | -0.20 | 0.29 | 28.73 |
| rs9436297_rs709158 | 8.02E-01 | 0.85 | 0.30 | 4.32E-03 | -0.06 | 0.30 | 26.64 |
| rs9436300_rs13099634 | 7.11E-01 | -1.17 | 0.41 | 4.13E-03 | -0.29 | 0.32 | 33.38 |
| rs9436742_rs2938395 | 8.53E-01 | 0.66 | 0.31 | 3.54E-02 | 0.04 | 0.32 | 13.71 |
| rs9871668_rs7188378 | 7.48E-01 | -0.48 | 0.21 | 2.50E-02 | 0.04 | 0.21 | 15.86 |
| rs9871668_rs741300 | 1.86E-01 | -0.69 | 0.34 | 4.47E-02 | -0.55 | 0.35 | 13.35 |
| rs9922619_rs7199363 | 2.35E-01 | -0.41 | 0.20 | 3.85E-02 | -0.30 | 0.21 | 13.47 |
| rs9925952_rs7184573 | 5.31E-01 | 0.69 | 0.32 | 3.13E-02 | -0.07 | 0.34 | 14.06 |
| rs9940629_rs12444954 | 6.91E-01 | 0.33 | 0.16 | 3.87E-02 | 0.10 | 0.16 | 12.90 |
| rs9940629_rs7186220 | 8.60E-02 | 0.38 | 0.14 | 8.43E-03 | 0.28 | 0.15 | 21.23 |
| rs9940629_rs9924983 | 9.24E-01 | 0.43 | 0.14 | 2.57E-03 | 0.03 | 0.14 | 28.24 |

Table 3s.7. LLFS white males.

| SNP | pPhenoSNP | beta_1 | SE_1 | pExpSNP | beta_2 | SE_2 | F |
|---|---|---|---|---|---|---|---|
| rs4468199_rs2167270 | 1.80E-01 | 0.94 | 0.29 | 3.47E-03 | 0.44 | 0.29 | 10.19 |
| rs4468199_rs9928094 | 4.81E-01 | 1.11 | 0.31 | 1.36E-03 | 0.27 | 0.31 | 12.65 |
| rs4468199_rs9930333 | 4.81E-01 | 1.11 | 0.31 | 1.36E-03 | 0.27 | 0.31 | 12.65 |
| rs4468199_rs9939973 | 4.81E-01 | 1.11 | 0.31 | 1.36E-03 | 0.27 | 0.31 | 12.65 |
| rs4468199_rs1421085 | 5.27E-01 | 1.16 | 0.32 | 1.23E-03 | 0.26 | 0.32 | 12.91 |
| rs4468199_rs1121980 | 4.81E-01 | 1.11 | 0.31 | 1.36E-03 | 0.27 | 0.31 | 12.65 |
| rs4468199_rs7193144 | 4.30E-01 | 1.22 | 0.33 | 8.28E-04 | 0.31 | 0.32 | 14.03 |
| rs4468199_rs8057044 | 5.87E-01 | 1.15 | 0.32 | 1.04E-03 | 0.22 | 0.31 | 13.40 |
| rs4468199_rs17817449 | 3.84E-01 | 1.25 | 0.33 | 6.76E-04 | 0.33 | 0.32 | 14.61 |
| rs4468199_rs8050136 | 3.84E-01 | 1.25 | 0.33 | 6.76E-04 | 0.33 | 0.32 | 14.61 |
| rs4468199_rs3751812 | 4.30E-01 | 1.22 | 0.33 | 8.28E-04 | 0.31 | 0.32 | 14.03 |
| rs4468199_rs9936385 | 4.30E-01 | 1.22 | 0.33 | 8.28E-04 | 0.31 | 0.32 | 14.03 |
| rs4468199_rs7185735 | 4.30E-01 | 1.22 | 0.33 | 8.28E-04 | 0.31 | 0.32 | 14.03 |
| rs4468199_rs17817964 | 4.30E-01 | 1.22 | 0.33 | 8.28E-04 | 0.31 | 0.32 | 14.03 |
| rs4468199_rs9922708 | 5.04E-01 | 1.12 | 0.30 | 9.73E-04 | 0.25 | 0.30 | 13.57 |
| rs4468199_rs9922619 | 5.04E-01 | 1.12 | 0.30 | 9.73E-04 | 0.25 | 0.30 | 13.57 |
| rs4468199_rs12149832 | 5.52E-01 | 1.16 | 0.31 | 8.92E-04 | 0.24 | 0.30 | 13.82 |
| rs4468199_rs7191718 | 8.21E-01 | 1.11 | 0.31 | 1.18E-03 | -0.01 | 0.32 | 13.04 |

| | | | | | | | |
|---|---|---|---|---|---|---|---|
| rs4468199_rs9924877 | 8.79E-01 | 1.16 | 0.35 | 2.64E-03 | 0.12 | 0.36 | 10.89 |
| rs4468199_rs918032 | 4.28E-01 | 1.23 | 0.31 | 4.63E-04 | -0.18 | 0.33 | 15.71 |
| rs4468199_rs1008400 | 3.49E-01 | 1.22 | 0.31 | 4.26E-04 | -0.23 | 0.33 | 15.96 |
| rs4468199_rs697769 | 4.68E-01 | 1.26 | 0.31 | 3.89E-04 | -0.16 | 0.33 | 16.23 |
| rs4468199_rs8053279 | 2.97E-01 | 0.99 | 0.30 | 2.85E-03 | 0.37 | 0.30 | 10.69 |
| rs4468199_rs12597715 | 2.04E-01 | 0.98 | 0.30 | 3.11E-03 | 0.43 | 0.30 | 10.47 |
| rs3790433_rs2167270 | 2.07E-01 | 0.78 | 0.25 | 3.67E-03 | 0.35 | 0.25 | 10.05 |
| rs3790433_rs9928094 | 3.58E-01 | 0.93 | 0.26 | 1.22E-03 | 0.27 | 0.25 | 12.95 |
| rs3790433_rs9930333 | 3.58E-01 | 0.93 | 0.26 | 1.22E-03 | 0.27 | 0.25 | 12.95 |
| rs3790433_rs9939973 | 3.58E-01 | 0.93 | 0.26 | 1.22E-03 | 0.27 | 0.25 | 12.95 |
| rs3790433_rs1421085 | 4.73E-01 | 0.95 | 0.26 | 1.06E-03 | 0.22 | 0.25 | 13.33 |
| rs3790433_rs1121980 | 3.58E-01 | 0.93 | 0.26 | 1.22E-03 | 0.27 | 0.25 | 12.95 |
| rs3790433_rs7193144 | 3.41E-01 | 1.00 | 0.26 | 6.12E-04 | 0.28 | 0.25 | 14.90 |
| rs3790433_rs8057044 | 4.10E-01 | 0.92 | 0.25 | 1.04E-03 | 0.24 | 0.24 | 13.38 |
| rs3790433_rs17817449 | 3.13E-01 | 1.01 | 0.26 | 5.36E-04 | 0.29 | 0.25 | 15.28 |
| rs3790433_rs8050136 | 3.13E-01 | 1.01 | 0.26 | 5.36E-04 | 0.29 | 0.25 | 15.28 |
| rs3790433_rs3751812 | 3.41E-01 | 1.00 | 0.26 | 6.12E-04 | 0.28 | 0.25 | 14.90 |
| rs3790433_rs9936385 | 3.41E-01 | 1.00 | 0.26 | 6.12E-04 | 0.28 | 0.25 | 14.90 |
| rs3790433_rs7185735 | 3.41E-01 | 1.00 | 0.26 | 6.12E-04 | 0.28 | 0.25 | 14.90 |
| rs3790433_rs17817964 | 3.41E-01 | 1.00 | 0.26 | 6.12E-04 | 0.28 | 0.25 | 14.90 |
| rs3790433_rs9922708 | 3.16E-01 | 0.97 | 0.25 | 6.49E-04 | 0.29 | 0.24 | 14.72 |
| rs3790433_rs9922619 | 3.16E-01 | 0.97 | 0.25 | 6.49E-04 | 0.29 | 0.24 | 14.72 |
| rs3790433_rs12149832 | 4.20E-01 | 0.99 | 0.25 | 5.67E-04 | 0.24 | 0.24 | 15.11 |
| rs3790433_rs11642841 | 4.43E-01 | 0.88 | 0.25 | 1.59E-03 | 0.23 | 0.25 | 12.22 |
| rs3790433_rs13337496 | 2.93E-01 | 1.10 | 0.32 | 1.89E-03 | 0.38 | 0.31 | 11.77 |
| rs3790433_rs73617837 | 2.41E-01 | 1.06 | 0.33 | 3.63E-03 | 0.43 | 0.33 | 10.08 |
| rs3790433_rs9936319 | 2.41E-01 | 1.06 | 0.33 | 3.63E-03 | 0.43 | 0.33 | 10.08 |
| rs3790433_rs77140256 | 2.41E-01 | 1.06 | 0.33 | 3.63E-03 | 0.43 | 0.33 | 10.08 |
| rs3790433_rs7191718 | 7.17E-01 | 0.83 | 0.24 | 1.73E-03 | 0.13 | 0.24 | 11.99 |
| rs3790433_rs697769 | 8.03E-01 | 0.93 | 0.26 | 1.12E-03 | -0.01 | 0.25 | 13.18 |
| rs77848204_rs7191718 | 9.79E-01 | 1.05 | 0.32 | 2.97E-03 | 0.07 | 0.34 | 10.58 |
| rs75465244_rs2167270 | 1.80E-01 | 0.94 | 0.29 | 3.47E-03 | 0.44 | 0.29 | 10.19 |
| rs75465244_rs9928094 | 4.81E-01 | 1.11 | 0.31 | 1.36E-03 | 0.27 | 0.31 | 12.65 |

| | | | | | | | |
|---|---|---|---|---|---|---|---|
| rs75465244_rs9930333 | 4.81E-01 | 1.11 | 0.31 | 1.36E-03 | 0.27 | 0.31 | 12.65 |
| rs75465244_rs9939973 | 4.81E-01 | 1.11 | 0.31 | 1.36E-03 | 0.27 | 0.31 | 12.65 |
| rs75465244_rs1421085 | 5.27E-01 | 1.16 | 0.32 | 1.23E-03 | 0.26 | 0.32 | 12.91 |
| rs75465244_rs1121980 | 4.81E-01 | 1.11 | 0.31 | 1.36E-03 | 0.27 | 0.31 | 12.65 |
| rs75465244_rs7193144 | 4.30E-01 | 1.22 | 0.33 | 8.28E-04 | 0.31 | 0.32 | 14.03 |
| rs75465244_rs8057044 | 5.87E-01 | 1.15 | 0.32 | 1.04E-03 | 0.22 | 0.31 | 13.40 |
| rs75465244_rs8050136 | 3.84E-01 | 1.25 | 0.33 | 6.76E-04 | 0.33 | 0.32 | 14.61 |
| rs75465244_rs3751812 | 4.30E-01 | 1.22 | 0.33 | 8.28E-04 | 0.31 | 0.32 | 14.03 |
| rs75465244_rs9936385 | 4.30E-01 | 1.22 | 0.33 | 8.28E-04 | 0.31 | 0.32 | 14.03 |
| rs75465244_rs7185735 | 4.30E-01 | 1.22 | 0.33 | 8.28E-04 | 0.31 | 0.32 | 14.03 |
| rs75465244_rs9922708 | 5.04E-01 | 1.12 | 0.30 | 9.73E-04 | 0.25 | 0.30 | 13.57 |
| rs75465244_rs9922619 | 5.04E-01 | 1.12 | 0.30 | 9.73E-04 | 0.25 | 0.30 | 13.57 |
| rs75465244_rs7191718 | 8.21E-01 | 1.11 | 0.31 | 1.18E-03 | -0.01 | 0.32 | 13.04 |
| rs75465244_rs9924877 | 8.79E-01 | 1.16 | 0.35 | 2.64E-03 | 0.12 | 0.36 | 10.89 |
| rs75465244_rs918032 | 4.28E-01 | 1.23 | 0.31 | 4.63E-04 | -0.18 | 0.33 | 15.71 |
| rs75465244_rs1008400 | 3.49E-01 | 1.22 | 0.31 | 4.26E-04 | -0.23 | 0.33 | 15.96 |
| rs75465244_rs697769 | 4.68E-01 | 1.26 | 0.31 | 3.89E-04 | -0.16 | 0.33 | 16.23 |
| rs75465244_rs8053279 | 2.97E-01 | 0.99 | 0.30 | 2.85E-03 | 0.37 | 0.30 | 10.69 |
| rs9436301_rs7193144 | 2.80E-01 | 0.85 | 0.25 | 2.12E-03 | 0.31 | 0.25 | 11.46 |
| rs9436301_rs8057044 | 3.09E-01 | 0.80 | 0.25 | 2.95E-03 | 0.29 | 0.24 | 10.60 |
| rs9436301_rs17817449 | 2.55E-01 | 0.86 | 0.25 | 1.84E-03 | 0.32 | 0.25 | 11.84 |
| rs9436301_rs8050136 | 2.55E-01 | 0.86 | 0.25 | 1.84E-03 | 0.32 | 0.25 | 11.84 |
| rs9436301_rs3751812 | 2.80E-01 | 0.85 | 0.25 | 2.12E-03 | 0.31 | 0.25 | 11.46 |
| rs9436301_rs9936385 | 2.80E-01 | 0.85 | 0.25 | 2.12E-03 | 0.31 | 0.25 | 11.46 |
| rs9436301_rs7185735 | 2.80E-01 | 0.85 | 0.25 | 2.12E-03 | 0.31 | 0.25 | 11.46 |
| rs9436301_rs17817964 | 2.80E-01 | 0.85 | 0.25 | 2.12E-03 | 0.31 | 0.25 | 11.46 |
| rs9436301_rs9922708 | 1.88E-01 | 0.82 | 0.25 | 2.57E-03 | 0.36 | 0.24 | 10.96 |
| rs9436301_rs9922619 | 1.88E-01 | 0.82 | 0.25 | 2.57E-03 | 0.36 | 0.24 | 10.96 |
| rs9436301_rs12149832 | 2.59E-01 | 0.83 | 0.25 | 2.23E-03 | 0.31 | 0.24 | 11.33 |
| rs9436301_rs7191718 | 8.54E-01 | 0.80 | 0.25 | 3.09E-03 | 0.09 | 0.25 | 10.49 |
| rs9436301_rs918032 | 9.44E-01 | 0.83 | 0.24 | 1.75E-03 | 0.06 | 0.25 | 11.97 |
| rs9436301_rs1008400 | 9.55E-01 | 0.83 | 0.24 | 1.61E-03 | 0.03 | 0.24 | 12.20 |
| rs9436301_rs697769 | 8.99E-01 | 0.85 | 0.24 | 1.54E-03 | 0.08 | 0.25 | 12.31 |

| | | | | | | | |
|---|---|---|---|---|---|---|---|
| rs9436302_rs697769 | 7.08E-01 | 0.90 | 0.25 | 1.44E-03 | 0.14 | 0.26 | 12.50 |
| rs1887285_rs2960422 | 9.29E-02 | 0.99 | 0.30 | 2.81E-03 | 0.55 | 0.30 | 10.73 |
| rs1887285_rs2167270 | 2.07E-01 | 0.99 | 0.29 | 2.23E-03 | 0.42 | 0.29 | 11.33 |
| rs1887285_rs9928094 | 5.45E-01 | 1.13 | 0.31 | 1.01E-03 | 0.24 | 0.30 | 13.46 |
| rs1887285_rs9930333 | 5.45E-01 | 1.13 | 0.31 | 1.01E-03 | 0.24 | 0.30 | 13.46 |
| rs1887285_rs9939973 | 5.45E-01 | 1.13 | 0.31 | 1.01E-03 | 0.24 | 0.30 | 13.46 |
| rs1887285_rs1421085 | 5.96E-01 | 1.17 | 0.32 | 9.27E-04 | 0.22 | 0.31 | 13.71 |
| rs1887285_rs1121980 | 5.45E-01 | 1.13 | 0.31 | 1.01E-03 | 0.24 | 0.30 | 13.46 |
| rs1887285_rs7193144 | 4.94E-01 | 1.24 | 0.32 | 6.32E-04 | 0.27 | 0.31 | 14.80 |
| rs1887285_rs8057044 | 6.56E-01 | 1.17 | 0.31 | 7.76E-04 | 0.19 | 0.30 | 14.21 |
| rs1887285_rs17817449 | 4.46E-01 | 1.26 | 0.32 | 5.20E-04 | 0.29 | 0.31 | 15.37 |
| rs1887285_rs8050136 | 4.46E-01 | 1.26 | 0.32 | 5.20E-04 | 0.29 | 0.31 | 15.37 |
| rs1887285_rs3751812 | 4.94E-01 | 1.24 | 0.32 | 6.32E-04 | 0.27 | 0.31 | 14.80 |
| rs1887285_rs9936385 | 4.94E-01 | 1.24 | 0.32 | 6.32E-04 | 0.27 | 0.31 | 14.80 |
| rs1887285_rs7185735 | 4.94E-01 | 1.24 | 0.32 | 6.32E-04 | 0.27 | 0.31 | 14.80 |
| rs1887285_rs17817964 | 4.94E-01 | 1.24 | 0.32 | 6.32E-04 | 0.27 | 0.31 | 14.80 |
| rs1887285_rs9922708 | 5.67E-01 | 1.13 | 0.30 | 7.33E-04 | 0.22 | 0.29 | 14.38 |
| rs1887285_rs9922619 | 5.67E-01 | 1.13 | 0.30 | 7.33E-04 | 0.22 | 0.29 | 14.38 |
| rs1887285_rs12149832 | 6.19E-01 | 1.17 | 0.31 | 6.77E-04 | 0.21 | 0.30 | 14.60 |
| rs1887285_rs7191718 | 7.52E-01 | 1.17 | 0.31 | 7.53E-04 | -0.03 | 0.32 | 14.30 |
| rs1887285_rs12933996 | 7.79E-01 | 1.03 | 0.32 | 3.30E-03 | -0.03 | 0.34 | 10.32 |
| rs1887285_rs9924877 | 9.62E-01 | 1.17 | 0.34 | 1.95E-03 | 0.08 | 0.35 | 11.68 |
| rs1887285_rs7206456 | 3.88E-01 | 1.16 | 0.36 | 3.00E-03 | -0.28 | 0.41 | 10.56 |
| rs1887285_rs918032 | 3.86E-01 | 1.25 | 0.31 | 3.49E-04 | -0.20 | 0.33 | 16.56 |
| rs1887285_rs1008400 | 3.15E-01 | 1.23 | 0.30 | 3.19E-04 | -0.24 | 0.32 | 16.83 |
| rs1887285_rs697769 | 4.23E-01 | 1.28 | 0.31 | 2.94E-04 | -0.18 | 0.33 | 17.08 |
| rs1887285_rs8053279 | 2.97E-01 | 0.99 | 0.30 | 2.85E-03 | 0.37 | 0.30 | 10.69 |
| rs1887285_rs12597715 | 2.04E-01 | 0.98 | 0.30 | 3.11E-03 | 0.43 | 0.30 | 10.47 |
| rs7602_rs697769 | 7.08E-01 | 0.90 | 0.25 | 1.44E-03 | 0.14 | 0.26 | 12.50 |
| rs1045895_rs35510800 | 9.02E-01 | -1.25 | 0.39 | 3.25E-03 | -0.08 | 0.29 | 10.36 |
| rs970468_rs13428113 | 9.17E-02 | -1.08 | 0.33 | 2.62E-03 | 0.38 | 0.26 | 10.91 |
| rs75169973_rs2167270 | 1.31E-01 | 1.12 | 0.34 | 2.57E-03 | 0.56 | 0.33 | 10.96 |
| rs11808888_rs2167270 | 3.99E-01 | 0.98 | 0.30 | 2.95E-03 | 0.31 | 0.31 | 10.61 |

| | | | | | | | |
|---|---|---|---|---|---|---|---|
| rs17127673_rs2167270 | 3.99E-01 | 0.98 | 0.30 | 2.95E-03 | 0.31 | 0.31 | 10.61 |
| rs17127677_rs697769 | 5.80E-01 | 1.04 | 0.28 | 8.92E-04 | -0.10 | 0.30 | 13.82 |
| rs6694528_rs2167270 | 3.99E-01 | 0.98 | 0.30 | 2.95E-03 | 0.31 | 0.31 | 10.61 |
| rs2767485_rs7191718 | 9.42E-01 | 0.84 | 0.26 | 2.91E-03 | 0.07 | 0.27 | 10.64 |
| rs2767485_rs697769 | 7.77E-01 | 0.95 | 0.26 | 1.00E-03 | -0.02 | 0.27 | 13.49 |
| rs1475397_rs4135294 | 1.21E-01 | 1.22 | 0.37 | 2.99E-03 | -0.66 | 0.47 | 10.57 |
| rs1475397_rs13099634 | 3.76E-01 | 1.06 | 0.32 | 2.66E-03 | -0.25 | 0.36 | 10.87 |
| rs1782755_rs2167270 | 2.44E-01 | 0.93 | 0.29 | 3.56E-03 | 0.40 | 0.30 | 10.13 |
| rs1782755_rs7191718 | 9.42E-01 | 0.84 | 0.26 | 2.91E-03 | 0.07 | 0.27 | 10.64 |
| rs1782755_rs16952987 | 3.46E-01 | 1.09 | 0.34 | 2.95E-03 | -0.29 | 0.38 | 10.61 |
| rs1171278_rs2167270 | 2.44E-01 | 0.93 | 0.29 | 3.56E-03 | 0.40 | 0.30 | 10.13 |
| rs1171278_rs7191718 | 9.42E-01 | 0.84 | 0.26 | 2.91E-03 | 0.07 | 0.27 | 10.64 |
| rs1171278_rs16952987 | 3.46E-01 | 1.09 | 0.34 | 2.95E-03 | -0.29 | 0.38 | 10.61 |
| rs1171281_rs2167270 | 2.44E-01 | 0.93 | 0.29 | 3.56E-03 | 0.40 | 0.30 | 10.13 |
| rs1171281_rs7191718 | 9.42E-01 | 0.84 | 0.26 | 2.91E-03 | 0.07 | 0.27 | 10.64 |
| rs1171281_rs16952987 | 3.46E-01 | 1.09 | 0.34 | 2.95E-03 | -0.29 | 0.38 | 10.61 |
| rs62164899_rs7191718 | 9.78E-01 | 1.16 | 0.34 | 2.07E-03 | 0.07 | 0.35 | 11.52 |
| rs6794024_rs7191718 | 3.83E-01 | 0.85 | 0.27 | 3.56E-03 | -0.20 | 0.29 | 10.13 |

Table 3s.8. LLFS white non-stratified.

| SNP | pPhenoSNP | beta_1 | SE_1 | pExpSNP | beta_2 | SE_2 | F |
|---|---|---|---|---|---|---|---|
| rs4468199_rs12629240 | 5.07E-01 | 0.91 | 0.28 | 1.69E-03 | 0.33 | 0.33 | 10.27 |
| rs4468199_rs9310401 | 5.07E-01 | 0.91 | 0.28 | 1.69E-03 | 0.33 | 0.33 | 10.27 |
| rs4468199_rs11715073 | 5.07E-01 | 0.91 | 0.28 | 1.69E-03 | 0.33 | 0.33 | 10.27 |
| rs4468199_rs11128598 | 5.07E-01 | 0.91 | 0.28 | 1.69E-03 | 0.33 | 0.33 | 10.27 |
| rs4468199_rs12496505 | 5.07E-01 | 0.91 | 0.28 | 1.69E-03 | 0.33 | 0.33 | 10.27 |
| rs4468199_rs12487012 | 5.07E-01 | 0.91 | 0.28 | 1.69E-03 | 0.33 | 0.33 | 10.27 |
| rs4468199_rs13070963 | 5.07E-01 | 0.91 | 0.28 | 1.69E-03 | 0.33 | 0.33 | 10.27 |
| rs4468199_rs12636454 | 5.07E-01 | 0.91 | 0.28 | 1.69E-03 | 0.33 | 0.33 | 10.27 |
| rs4468199_rs11128599 | 4.52E-01 | 0.96 | 0.29 | 1.19E-03 | 0.36 | 0.33 | 10.97 |
| rs4468199_rs12493718 | 5.07E-01 | 0.91 | 0.28 | 1.69E-03 | 0.33 | 0.33 | 10.27 |
| rs4468199_rs12496005 | 5.07E-01 | 0.91 | 0.28 | 1.69E-03 | 0.33 | 0.33 | 10.27 |

| | | | | | | | |
|---|---|---|---|---|---|---|---|
| rs4468199_rs4145574 | 5.07E-01 | 0.91 | 0.28 | 1.69E-03 | 0.33 | 0.33 | 10.27 |
| rs4468199_rs4145573 | 5.07E-01 | 0.91 | 0.28 | 1.69E-03 | 0.33 | 0.33 | 10.27 |
| rs4468199_rs2028760 | 5.07E-01 | 0.91 | 0.28 | 1.69E-03 | 0.33 | 0.33 | 10.27 |
| rs4468199_rs1122648 | 5.07E-01 | 0.91 | 0.28 | 1.69E-03 | 0.33 | 0.33 | 10.27 |
| rs4468199_rs11128601 | 5.07E-01 | 0.91 | 0.28 | 1.69E-03 | 0.33 | 0.33 | 10.27 |
| rs75465244_rs9310401 | 5.07E-01 | 0.91 | 0.28 | 1.69E-03 | 0.33 | 0.33 | 10.27 |
| rs75465244_rs4145574 | 5.07E-01 | 0.91 | 0.28 | 1.69E-03 | 0.33 | 0.33 | 10.27 |
| rs75465244_rs4145573 | 5.07E-01 | 0.91 | 0.28 | 1.69E-03 | 0.33 | 0.33 | 10.27 |
| rs75465244_rs2028760 | 5.07E-01 | 0.91 | 0.28 | 1.69E-03 | 0.33 | 0.33 | 10.27 |
| rs75465244_rs1122648 | 5.07E-01 | 0.91 | 0.28 | 1.69E-03 | 0.33 | 0.33 | 10.27 |
| rs1887285_rs6768587 | 6.68E-01 | 0.88 | 0.27 | 1.48E-03 | 0.25 | 0.31 | 10.52 |
| rs1887285_rs9817428 | 6.68E-01 | 0.88 | 0.27 | 1.48E-03 | 0.25 | 0.31 | 10.52 |
| rs1887285_rs12631028 | 6.68E-01 | 0.88 | 0.27 | 1.48E-03 | 0.25 | 0.31 | 10.52 |
| rs1887285_rs12636461 | 6.68E-01 | 0.88 | 0.27 | 1.48E-03 | 0.25 | 0.31 | 10.52 |
| rs1887285_rs13076055 | 6.68E-01 | 0.88 | 0.27 | 1.48E-03 | 0.25 | 0.31 | 10.52 |
| rs1887285_rs11128596 | 6.68E-01 | 0.88 | 0.27 | 1.48E-03 | 0.25 | 0.31 | 10.52 |
| rs1887285_rs11128597 | 6.68E-01 | 0.88 | 0.27 | 1.48E-03 | 0.25 | 0.31 | 10.52 |
| rs1887285_rs17036242 | 6.68E-01 | 0.88 | 0.27 | 1.48E-03 | 0.25 | 0.31 | 10.52 |
| rs1887285_rs12629240 | 6.10E-01 | 0.92 | 0.28 | 1.07E-03 | 0.27 | 0.31 | 11.18 |
| rs1887285_rs9310401 | 6.10E-01 | 0.92 | 0.28 | 1.07E-03 | 0.27 | 0.31 | 11.18 |
| rs1887285_rs11715073 | 6.10E-01 | 0.92 | 0.28 | 1.07E-03 | 0.27 | 0.31 | 11.18 |
| rs1887285_rs11128598 | 6.10E-01 | 0.92 | 0.28 | 1.07E-03 | 0.27 | 0.31 | 11.18 |
| rs1887285_rs12496505 | 6.10E-01 | 0.92 | 0.28 | 1.07E-03 | 0.27 | 0.31 | 11.18 |
| rs1887285_rs12487012 | 6.10E-01 | 0.92 | 0.28 | 1.07E-03 | 0.27 | 0.31 | 11.18 |
| rs1887285_rs13070963 | 6.10E-01 | 0.92 | 0.28 | 1.07E-03 | 0.27 | 0.31 | 11.18 |
| rs1887285_rs12636454 | 6.10E-01 | 0.92 | 0.28 | 1.07E-03 | 0.27 | 0.31 | 11.18 |
| rs1887285_rs11128599 | 5.53E-01 | 0.96 | 0.28 | 7.67E-04 | 0.29 | 0.31 | 11.85 |
| rs1887285_rs12493718 | 6.10E-01 | 0.92 | 0.28 | 1.07E-03 | 0.27 | 0.31 | 11.18 |
| rs1887285_rs12496005 | 6.10E-01 | 0.92 | 0.28 | 1.07E-03 | 0.27 | 0.31 | 11.18 |
| rs1887285_rs4145574 | 6.10E-01 | 0.92 | 0.28 | 1.07E-03 | 0.27 | 0.31 | 11.18 |
| rs1887285_rs4145573 | 6.10E-01 | 0.92 | 0.28 | 1.07E-03 | 0.27 | 0.31 | 11.18 |
| rs1887285_rs2028760 | 6.10E-01 | 0.92 | 0.28 | 1.07E-03 | 0.27 | 0.31 | 11.18 |
| rs1887285_rs1122648 | 6.10E-01 | 0.92 | 0.28 | 1.07E-03 | 0.27 | 0.31 | 11.18 |

| | | | | | | | |
|---|---|---|---|---|---|---|---|
| rs1887285_rs11128601 | 6.10E-01 | 0.92 | 0.28 | 1.07E-03 | 0.27 | 0.31 | 11.18 |
| rs75169973_rs741300 | 3.57E-01 | 0.75 | 0.22 | 9.66E-04 | -0.21 | 0.31 | 11.38 |
| rs75169973_rs708278 | 1.13E-01 | 0.77 | 0.24 | 1.59E-03 | -0.51 | 0.38 | 10.39 |

Table 3s.9. LLFS white females (using original SNPs selected from GWAS).

| SNP | pPhenoSNP | beta_1 | SE_1 | pExpSNP | beta_2 | SE_2 | F |
|---|---|---|---|---|---|---|---|
| rs10881900 | 8.33E-01 | 0.76 | 0.21 | 6.43E-04 | -0.23 | 0.13 | 13.16 |
| rs4671402 | 9.30E-01 | -0.69 | 0.20 | 1.34E-03 | 0.22 | 0.13 | 11.46 |
| rs7120582 | 1.50E-01 | -0.83 | 0.24 | 9.50E-04 | -0.20 | 0.15 | 12.23 |
| rs76749554 | 9.66E-01 | -0.84 | 0.25 | 1.39E-03 | -0.28 | 0.15 | 11.39 |

Table 3s.10. LLFS white males (using original SNPs selected from GWAS).

| SNP | pPhenoSNP | beta_1 | SE_1 | pExpSNP | beta_2 | SE_2 | F |
|---|---|---|---|---|---|---|---|
| rs11689002 | 8.50E-01 | -1.01 | 0.30 | 1.84E-03 | 0.18 | 0.13 | 11.17 |
| rs12462704 | 5.32E-01 | -0.86 | 0.27 | 2.92E-03 | -0.19 | 0.13 | 10.08 |
| rs517711 | 6.62E-01 | 1.03 | 0.31 | 2.12E-03 | 0.28 | 0.16 | 10.83 |
| rs4237817 | 5.34E-02 | 0.82 | 0.25 | 2.07E-03 | 0.23 | 0.12 | 10.89 |
| rs36118060 | 4.62E-01 | 1.12 | 0.34 | 1.84E-03 | 0.22 | 0.16 | 11.17 |
| rs66990209 | 2.42E-01 | -0.92 | 0.26 | 1.21E-03 | -0.19 | 0.12 | 12.24 |
| rs72898727 | 7.33E-02 | 1.37 | 0.43 | 2.70E-03 | -0.60 | 0.25 | 10.26 |
| rs77238796 | 4.83E-01 | 1.47 | 0.44 | 2.01E-03 | 0.35 | 0.22 | 10.96 |
| rs7982809 | 7.85E-01 | 0.94 | 0.29 | 2.42E-03 | -0.18 | 0.15 | 10.52 |

Table 3s.11. LLFS white non-stratified (using original SNPs selected from GWAS).

| SNP | pPhenoSNP | beta_1 | SE_1 | pExpSNP | beta_2 | SE_2 | F |
|---|---|---|---|---|---|---|---|
| rs115500306 | 9.54E-01 | 1.08 | 0.34 | 1.58E-03 | -0.26 | 0.21 | 10.32 |
| rs11127167 | 1.29E-01 | 0.53 | 0.15 | 7.08E-04 | -0.13 | 0.09 | 11.90 |
| rs12422418 | 6.96E-01 | 1.14 | 0.26 | 2.24E-05 | 0.21 | 0.14 | 19.02 |
| rs17768871 | 2.77E-01 | 0.93 | 0.28 | 1.08E-03 | -0.44 | 0.20 | 11.06 |
| rs17512884 | 7.21E-02 | 0.96 | 0.29 | 1.20E-03 | -0.27 | 0.18 | 10.86 |
| rs1932955 | 8.96E-02 | 0.50 | 0.14 | 7.18E-04 | -0.13 | 0.09 | 11.87 |
| rs201626 | 6.72E-01 | 0.82 | 0.25 | 1.19E-03 | -0.22 | 0.16 | 10.87 |

| | | | | | | | |
|---|---|---|---|---|---|---|---|
| rs6876057 | 5.06E-01 | 1.06 | 0.30 | 5.05E-04 | -0.25 | 0.19 | 12.57 |
| rs7258465 | 9.12E-01 | 0.58 | 0.15 | 1.79E-04 | 0.12 | 0.09 | 14.68 |
| rs73852555 | 4.01E-01 | -0.61 | 0.19 | 1.23E-03 | 0.22 | 0.10 | 10.80 |
| rs9304382 | 1.96E-01 | 0.61 | 0.19 | 1.18E-03 | 0.15 | 0.12 | 10.89 |

**Table 4s**. MR analysis result in LLFS data using SNPs that are significantly associated with exposure variable (<0.05) from GWAS result and satisfied all IV tests

| strata | No. of Indiv. | No of IVs | IVW Estimate (95% CI) | IVW P value | Penalized IVW Estimate (95% CI) | Penalized IVW P value | Weighted Median Estimate (95% CI) | Weighted Median P value | MR-Egger Regression Intercept (95%CI) | E-value Average(min, max) |
|---|---|---|---|---|---|---|---|---|---|---|
| White female | 1064 | 4 | -0.014 (-0.348,0.319) | 9.323E-1 | -0.014 (-0.348,0.195) | 9.323E-1 | -0.028 (-0.256,0.200) | 0.812 | -2.575 (-4.198, -0.651) | 3.81 (3.40,4.08) |
| White male | 940 | 9 | 0.084 (-0.075,0.244) | 2.993E-1 | 0.156 (0.049,0.271) | **7.905E-3** | 0.205 (0.058,0.351) | **6.203E-3** | 0.386 (-0.609,1.381) | 5.35 (3.98,8.16) |
| White F+M | 2004 | 11 | -0.116 (-0.266,0.035) | 1.322E-1 | -0.200 (-0.328, -0.072) | **2.142E-3** | -0.241 (-0.402, -0.080) | **3.401E-3** | -0.023 (-0.408, 0.362) | 4.00 (2.67,5.70) |

**Figure 1s**. Top SNP-SNP interaction from obesity/BMI related genes that are significantly associated with overweight in HRS data.

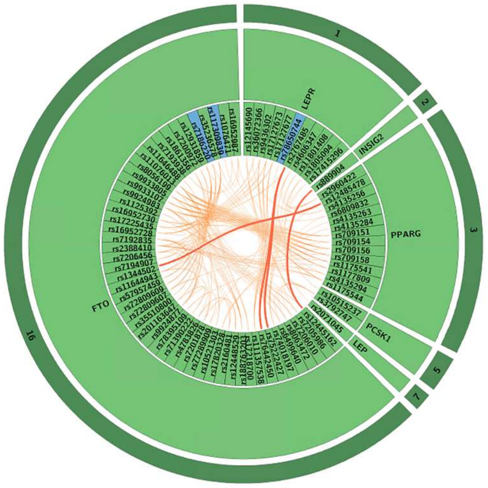


Notes: the curves connecting SNP pairs denote the effect of the SNP pairs on dichotomized BMI exposure variable (group 1: overweight, at ages [75,85]; group 0: normal weight, at ages [75,85]). The colors denote the p values of logistic regression: red < 10e-4, 10e-4 < orange < 10e-3; Single SNP p-values: blue < 0.05, green >0.05.

**Figure 2s**. Kaplan-Meier estimates of conditional survival functions in HRS data stratified by sex and race.

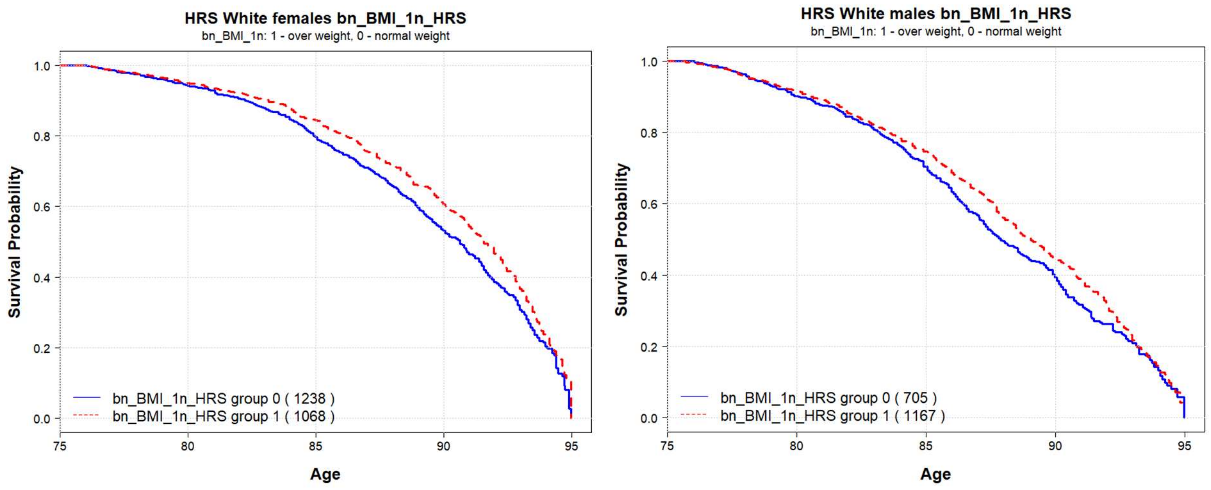

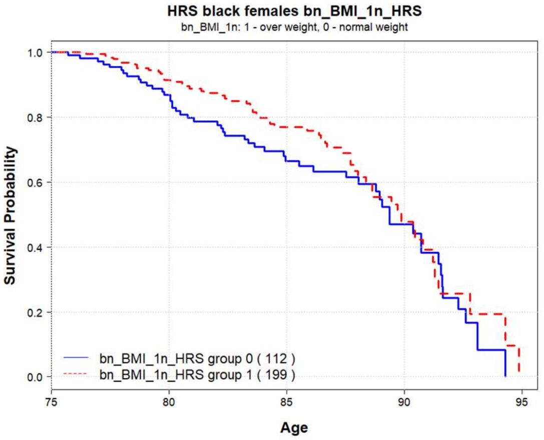


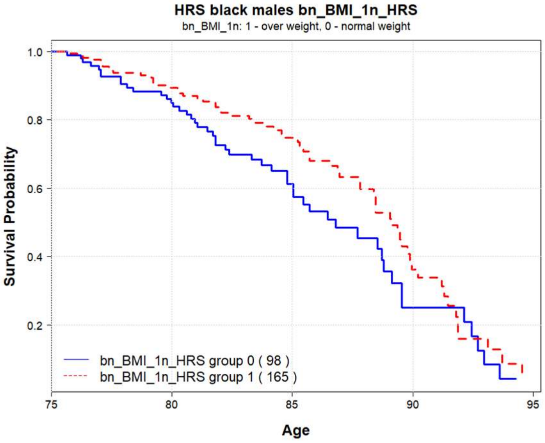


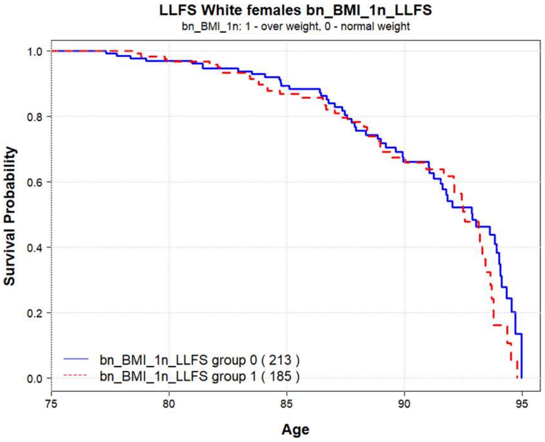


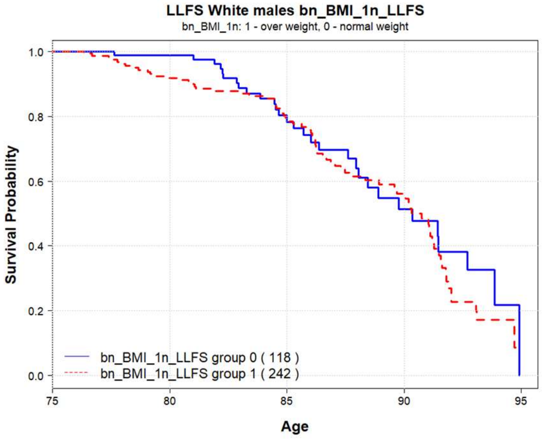


Notes: **bn_BMI_1n** denotes the name of the exposure binary variable (group 1: average BMI ≥ 25 and < 30 (“overweight”) at ages [75,85]; group 0: average BMI ≥ 18.5 and < 25 (“normal weight”) at ages [75,85]). Number of people in the groups are shown in the parentheses.
